# Does music support cognitive control and affective responses during acute exercise? An exploratory systematic review and meta-analysis

**DOI:** 10.1101/2025.01.28.25321259

**Authors:** Andrew Danso, Julia Vigl, Friederike Koehler, Keegan Knittle, Joshua S. Bamford, Patti Nijhuis, Eero A. Haapala, Ming Yu Claudia Wong, Shannon E. Wright, Margarida Baltazar, Nora Serres, Niels Chr. Hansen, Andrea Schiavio, Suvi Saarikallio, Geoff Luck

**Affiliations:** Centre of Excellence in Music, Mind, Body and Brain; Department of Music, Arts and Culture Studies, University of Jyväskylä, Finland; Department of Psychology, University of Innsbruck; Faculty of Sport and Health Sciences, University of Jyväskylä, Finland; Centre for the Study of Social Cohesion, University of Oxford, United Kingdom; Sports and Exercise Medicine, Faculty of Sport and Health Sciences, University of Jyväskylä, Finland; Institute of Biomedicine, School of Medicine, University of Eastern Finland, Finland; Children’s Health and Exercise Research Centre, Faculty of Health and Life Sciences, University of Exeter, United Kingdom; Department of Health and Physical Education, The Education University of Hong Kong, Hong Kong, China; Department of Psychology, University of the Fraser Valley, Canada; RITMO Centre for Interdisciplinary Studies in Rhythm, Time and Motion, University of Oslo; Royal Academy of Music Aarhus, Denmark; Interacting Minds Centre, School of Culture and Society, Denmark; School of Arts and Creative Technologies, University of York, United Kingdom

**Keywords:** music, exercise, cognitive control, attention allocation, inhibitory control, affect, systematic review

## Abstract

Cognitive control, defined as the allocation of mental resources required for goal-directed behaviour, is crucial for exercise participation as it is involved in regulating negative cognitive and affective responses caused by the demands of exercise. Research on both music and acute exercise separately show engagement of cognitive control processes and affective responses, with low-to-moderate exercise intensities reliably influencing cognitive and affective outcomes (e.g., core affect). However, the combined effects of music and acute exercise on cognitive control and affective outcomes remain underexplored. Accordingly, this review and meta-analysis explores how music influences cognitive control and affective outcomes during acute exercise. 10 studies met the inclusion criteria, with nine providing data for effect size calculations across 21 intervention arms. Meta-analyses revealed significant effects of music on attention allocation (*g* = 1.05, 95% CI [0.03, 2.07]; *p* = 0.04), inhibitory control (*g* = 1.87, 95% CI [0.37, 3.37]; *p* = 0.01), and core affect (*g* = 0.86, 95% CI [0.24, 1.48]; *p* < 0.01). Exercise intensity significantly moderated outcomes (*p* = 0.036), suggesting that higher intensities diminish the effectiveness of music in elevating cognitive control and affective outcomes during acute exercise. Findings were limited by high heterogeneity (*I²* > 97%) across study protocols and outcome measures. Due to the aforementioned heterogeneity, the findings of this review must be interpreted cautiously.

## Introduction

Cognitive control can be defined as the allocation of mental resources required for goal-directed behaviour (e.g., maintaining a steady pace during a long-distance run despite physical or mental fatigue) [1], and refers to the broader regulation of attention, thought, and action to accomplish goal-directed behavior. This is distinct from executive control, which specifically refers to the higher-order processes involved in decision-making to execute a task, such as the executive control of attention [1,2]. Cognitive control has implications for the degree to which recreationally active individuals manage their affective responses (e.g., pleasure) and cognitive demands (e.g., the allocation of attentional resources to maintain optimal movement patterns) during exercise [3,4]. Most of the literature has examined the psychological and psychophysiological effects of music in the exercise domain (see e.g., [3]), or addressed the differential effects of music and exercise on cognitive performance independently (see e.g., [5]); little attention has been given to the interplay between music and acute exercise influence on both cognitive control and affective domains.

In the context of exercise, cognitive control is crucial for exercise adherence [6] as it is involved in regulating negative cognitive and affective responses caused by the physical demands of exercise [3]. In addition, music listening during exercise has been shown to modulate cognitive and affective responses by influencing attention allocation through mechanisms such as auditory-motor coupling, where rhythmic synchronisation aligns movement with musical tempo [7,8]. Music listening during exercise may also influence attention allocation by directing focus either toward internal sensations, such as association, or external stimuli, such as dissociation. This can improve cognitive resource allocation while elevating affective responses during exercise, helping to offset interoceptive cues such as perceived exertion [3,9].

Cognitive control processes are activated through the dynamic interplay of attentional functions, such as alerting, orienting, and executive control, alongside broader cognitive processes that support goal-directed behaviour [1,2]. Cognitive control encompasses interrelated functions such as cognitive flexibility (shifting attention between internal and external stimuli), inhibitory control (suppressing impulses), and working memory (maintaining and manipulating information) [1,2,10]. Rather than a strictly hierarchical system, cognitive control entails both unity—shared mechanisms supporting goal-directed action—and diversity (e.g., the distinct functions that each component, such as inhibitory control, and attention allocation contributes to goal-directed behaviour), with distinct roles for its components [1,10].

Acute exercise, defined as a single bout of physical activity [11], has demonstrated immediate (e.g., measured immediately after the acute exercise session) elevated effects on cognitive flexibility and inhibitory control across diverse populations, particularly when performed at low-to-moderate intensities [9,12]. Similarly, there is a growing body of evidence showing the use of music at low-to-moderate exercise intensities significantly influencing cognitive control domains by minimising interoceptive demands and elevating affective responses [3,13,14]. However, these effects appear to diminish at higher exercise intensities, where cognitive and physiological demands increase, highlighting the need to clarify the contextual factors that modulate these outcomes.

Given the aforementioned cognitive benefits of acute exercise and music, coupled with the likelihood of these being perceived as making the exercise experience more pleasant, there has been growing interest in the application of music and acute exercise as a means to improve cognitive functioning [15]. The last decade has seen the emergence of an extensive corpus of work addressing the combined effects of music and acute exercise on cognitive control processes and affective outcomes (e.g., [13,14,16]). Accordingly, the present review explores the combined effects of music and acute exercise on cognitive control processes and affective outcomes by synthesising findings from meta-analyses and narrative syntheses. Specifically, it aims to answer two key research questions: (1) How does music listening influence cognitive control processes and affective responses during acute exercise? (2) Which cognitive control processes are most consistently influenced by music during exercise?

### The effects of physical exercise on cognitive control processes

Physical exercise has demonstrated significant effects on cognitive control processes, with acute and chronic forms yielding distinct effects (e.g., acute exercise improving inhibitory control, such as resisting distractions immediately post-exercise, and chronic exercise improving long-term working memory). While acute exercise refers to single bouts of physical activity, chronic exercise refers to a pattern of regular and repeated exercise over an extended period (e.g., weeks, months, years) [17]. Chronic exercise has been shown to provide sustained improvements in working memory and attention allocation [18], likely due to long-term neurocognitive adaptations induced by repeated exercise engagement [19]. In their examination of cognitive control processes such as task switching and cognitive flexibility, researchers [20–22] found that physical exercise elicited differential effects on these processes. Importantly, low-to-moderate exercise intensities (30% to 60% of VO2 max) were identified as a significant moderator of positive cognitive outcomes.

Exercise performed at low-to-moderate intensities has been consistently associated with improvements in cognitive control processes and affective responses [23]. In contrast, exercise performed at higher intensities (above 70% of VO2 max) often yields more variable cognitive effects [20,24], potentially due to individual differences in physiological and cognitive load (e.g., one’s tolerance for increased physical demands by regulating interoceptive processing during exercise) as well as affective responses at higher physical exertion levels.

A salient explanation is Dual-Mode Theory (DMT) suggesting higher exercise intensities increase variability in cognitive and affective responses due to shifts between automatic (e.g., Type 1, immediate responses of pleasure or discomfort) and controlled processes (e.g., Type 2, deliberate behaviours, such as adhering to an exercise routine based on long-term goals) [19,23,26]. DMT highlights the importance of the ventilatory threshold (VT) in determining affective and physiological responses. At moderate exercise intensities (up to VT), affective valence typically improves due to manageable physiological demands and the ability to process external stimuli (e.g., music). However, at higher intensities— beyond the respiratory compensation point (RCP)—affective responses become more variable, with declines in valence linked to the awareness of homeostatic disruptions [7]. This variability may impair cognitive outcomes by increasing cognitive load and diverting resources, which can lead to reduced inhibitory control when interoceptive demands are heightened. In contrast, low-to-moderate exercise intensities are associated with consistent cognitive-affective responses, providing favourable conditions for maintaining cognitive-affective responses during exercise.

### The role of music on cognitive and affective processes during exercise

Listening to music influences cognitive and affective responses during physical exercise by regulating affective responses, such as elevating affective valence during low-to-moderate cycling, diverting attention from psychophysical sensations by reducing perceived physical exertion [3], and fostering temporal prediction and rhythmic synchronisation [27]. These mechanisms align with the framework of DMT, where music interacts with exercise intensity to modulate affective and cognitive outcomes [26]. Rhythmic synchronisation, a form of auditory-motor coupling, aligns movement with musical tempo, supporting attention allocation and elevating affective responses during exercise [3,26]. Such effects are most pronounced within the DMT zone of response variability, where music interacts with individual preferences and physiological rhythms to influence the exercise experience [26].

Differences in music protocols, particularly the use of self-selected or researcher-selected music have been shown to influence cognitive processes. Recent work [28] demonstrated that participant-selected (self-selected) slow-tempo music elevated affective responses by reducing mental demand and frustration during tasks. Furthermore, individual fitness levels (e.g., trained athletes or those regularly engaging in aerobic exercise) moderate these effects, as fitter individuals may tolerate higher intensities and mitigate negative affective responses at such heightened exercise intensities [7]. Together, these factors— affective responses, auditory-motor coupling, and exercise intensity—are central to the extent to which cognitive resources can be directed toward cognitive control processes during physical exercise.

Music’s affective properties can be understood through the lens of the Circumplex Model of Affect [29,30], which frames core affect—defined as the fundamental neurophysiological state underlying emotional experiences—along two dimensions: affective valence (pleasant-unpleasant) and affective arousal (low-high energy). These dimensions are influenced by music as it is listened to during physical exercise. Specifically, moderate-to-fast-tempo (120–130 beats per minute, BPM), preference-based (e.g., self-selected) music may engender elevated affective valence and/or affective arousal, making it suitable for high-energy exercise (e.g., cycling at high intensities; see [31]). Conversely, preference-based slower-tempo music (60–90 BPM) may reduce affective arousal while maintaining elevated affective valence, thereby eliciting recuperation during low-intensity activities (e.g., walking or light cycling as a form of post-exercise recovery; [3]).

### The present study

Despite the meta-analysis by [5] demonstrating small but consistent positive effects of acute exercise on cognitive performance, particularly at low-to-moderate intensities, the interplay between music and acute exercise remains underexplored. While [5] primarily focused on exercise-cognitive paradigms such as examining moderate-intensity exercise with post-exercise Stroop task assessments, and moderators, such as fitness levels, they did not address the potential additive or interactive effects of auditory stimuli such as music. Thus, this review explores how music and acute exercise interact to influence cognitive control processes and affective outcomes. Understanding this interplay in a systematic manner is enticing, as both exercise (acute and chronic) and music are widely researched modalities that significantly influence cognitive and affective responses, thereby contributing to developing participant health and well-being interventions [15,32]. Accordingly, this review explores how music influences cognitive control processes and affective outcomes during acute exercise, potentially driven by mechanisms such as auditory-motor coupling, rhythmic synchronisation, and dissociation—effects that are most pronounced at low-to-moderate exercise intensities (e.g., [26,33]).

The central research questions guiding this review are:

*RQ1. How does music listening influence cognitive control processes and affective responses during acute exercise?*

*RQ2. Which cognitive control processes are most consistently affected by music during exercise?*

## Methods

In this systematic review, the primary comparison was between acute exercise with music listening compared to acute exercise without music listening in physically active adults aged 18 and over. This study was conducted and is reported in line with the Preferred Reporting Items for Systematic Reviews and Meta-Analyses (PRISMA) protocol [34].

### Eligibility criteria

We included studies that met the following criteria: The population comprised participants from various groups, including males and females across different age ranges, with a focus on acute exercise. The intervention involved music listening during exercise, targeting at least one cognitive control process of interest, such as attention allocation, inhibitory control, cognitive flexibility, working memory, or overall cognitive performance, and affective outcomes such as affect. The comparator condition was acute exercise alone, without music. Exceptions were made for studies where specific conditions (e.g., slower or faster mismatched music) could reasonably serve as proxies for a no-music control. The outcomes assessed were music and acute exercise effects on cognitive control processes and affective outcomes, specifically those listed above. The study design was limited to experimental designs, including randomised controlled trials (RCTs). Additionally, we included only articles published in English in peer-reviewed journals or as doctoral theses/dissertations, given the rigorous internal review processes associated with these formats.

### Information sources

We searched the following databases: (1) Web of Science, (2) SPORTDiscus, (3) MEDLINE, (4) Embase, (5) PubMed, (6) CINAHL Complete, (7) Cochrane Library, (8) Scopus databases, (9) PsycINFO, and (10) Google Scholar. The database search was supplemented by forward and backward snowball searches. The snowball search continued until no new sources could be identified. Specifically, the backward snowball search involved scanning the reference lists of all included articles for potential sources, while the forward snowball search identified additional studies by examining articles that cited the included studies. The initial inter-rater agreement for the identification of relevant sources was *k* = 0.88, indicating a strong level of agreement among the two individuals performing two independent snowball searches (AD, JV).

### Search strategy

A literature search was performed using terminology related to cognitive control processes being affected by music listening in physically active adults during exercise sessions. Specifically, the following search term was used: (TS=(“Cognitive control” OR “Executive functio*” OR “Inhibitory control” OR “Working memory” OR “Cognitive flexibility” OR “Task switching” OR “Attention” OR “Neurocognitive task*” OR “Goal-driven decision-making” OR “Dual-process theor*” OR “Autonomous processing” OR “Controlled processing” OR “Ironic process* theory” OR “Affect”)) AND (TS=(“Music listening” OR “Music intervention*” OR “Music-based intervention” OR “Music and cognition”)) AND (TS=(“Physical exercise” OR “Sports performance” OR “Exercise-induced cognition” OR “Performance enhance*”)). The full search strategy can be found in the review registration document (<u>OSF</u> https://osf.io/6eba5).

### Selection process and data collection process

The citations of all retrieved articles were imported into Zotero and all duplicates were manually removed. Study title and abstract were then screened by three authors (AD, JV, JB) using ASREVIEW [35]. If the article could not be excluded on the basis of the title or abstract, the retrieved full-text articles were then assessed for inclusion by two authors independently. At each stage, disagreements were discussed with a third author in cases where a consensus could not be achieved amongst the two initial screeners.

### Data extraction

The studies’ information was extracted to a spreadsheet. This included study characteristics, such as the targeted cognitive control process(s), the study design, cognitive process measurements, the music protocol, and study outcomes. Where available, quantitative data suitable for meta-analysis (e.g., mean values, standard deviations, or effect sizes for cognitive control outcomes) were also extracted. In instances where necessary data for meta analysis were not reported (*n*=2), corresponding study authors were contacted to obtain the missing data for analysis. If we did not receive the data required for meta-analysis (*n*=3), a narrative synthesis was conducted on those studies.

### Study risk of bias assessment

The quality of the studies were assessed by two authors (AD, JV) using the Joanna Briggs Institute critical appraisal checklist [JBI, 36], including tools for Quasi-Experimental Appraisal, and the Revised Checklist for RCTs (Figure 2). The JBI critical appraisal tools were chosen for their adaptability to diverse study designs, providing a structured and standardised approach to assessing risk of bias. While alternative tools such as the Cochrane framework are commonly used in clinical reviews, the JBI tools were more suitable for the interdisciplinary nature of this review, encompassing experimental studies in sports and cognitive psychology.

### Operationalisation of outcomes of interest

The main outcomes of interest in the review are operationalised in the following table (Table 1). It contains the cognitive process as the outcome of interest (see “outcome”), an “operational definition” of it, the “measurement tool(s)” and “key references” associated with the operationalisation (see e.g., Appendix A for a full description of the operationalisation of outcomes of interest in this review).

**Table 1.** Operationalisations of outcomes of interest.

| Outcome | Operational Definition | Measurement Tool(s) | Key References |
| --- | --- | --- | --- |
| Attention Allocation | The process of directing focus towards internal sensations (association) and/or external stimuli (dissociation) during physical exercise. | Association-Dissociation Questionnaire; dual-task paradigms | [37,38] |
| Inhibitory Control | Suppressing impulsive actions (e.g., abrupt changes in intensity or technique) and resisting distractions from internal (e.g., negative thoughts) and external (e.g., environmental factors) sources. It supports deliberate decision-making and attention allocation during physical exercise | Stroop Task, Go/No-Go Task | [1] |
| Cognitive Flexibility | Shifting attention between internal and external stimuli, modifying exercise behaviour based on feedback. | Cognitive Flexibility Inventory; task-switching paradigms | [1] |
| Task Switching | A component of cognitive flexibility involving attention allocation, to enable shifts between different cognitive and motor tasks. Efficiency and speed of task switching are influenced by task complexity, individual differences, and practice. | Task-switching paradigms; dual-task tests | [1] |
| Working Memory | Actively holds, manipulates, and processes information necessary for tasks like retaining instructions, monitoring feedback, and adapting to exercise behaviour. | N-back tasks; digit span tasks | [39] |
| Overall Cognitive Performance | Broadly the ability to process and respond to information during exercise. | Mini-Mental State Examination (MMSE); cognitive load measures | [39] |
| Core Affect | Encompasses both emotions and moods, characterised by two dimensions: valence (pleasure-displeasure) and arousal (activation-sleepiness). Affect arises from physiological processes, cognitive appraisals, and situational influences and can range from basic reflexive responses to complex emotions. | Feeling Scale, Felt Arousal Scale Two-Dimensional Mood Scale | [29,30] |

### Data synthesis and analysis methods

We conducted a narrative synthesis to qualitatively analyse findings across four identified cognitive processes: attention allocation, inhibitory control, cognitive flexibility and working memory. All affective outcomes were analysed under core affect. Studies were categorised by their primary outcomes, and trends were identified by examining the direction and magnitude of effects, alongside variations in study characteristics, such as participant demographics, exercise intensity, music conditions, and methodological designs. Heterogeneity was addressed by highlighting contextual factors, including differences in exercise protocols (e.g., exercise intensity, duration, modality, task type) and music conditions (e.g., tempo-matched, self-selected music).

To examine how music listening may exert an effect on cognitive control process outcomes, we performed a meta-analysis of the outcomes of interest in eligible studies. Eligible studies included those that compared music listening with acute exercise, to acute exercise without music listening, with both a control group and an intervention arm targeting the outcomes of interest. Additionally, at least three studies needed to report suitable, quantifiable data to be included in a single meta-analytic cluster.

For quantitative synthesis, we calculated Hedges’ *g* effect sizes and standard errors using the tool developed by [40]. In cases where multiple data points were extracted for the same outcome, weighted averages were calculated in Microsoft Excel to standardise Hedges’ *g* and standard error. These standardised data were then imported into JASP [41], where separate random-effects meta-analyses using the inverse-variance weighting method [DerSimonian & Laird method, [42] were conducted for the outcomes attention allocation, inhibitory control, and core affect. For the remaining outcomes, insufficient data were available to meet this criterion. Heterogeneity was assessed using the *I²* statistic, *τ²* statistic, and Cochran’s *Q* statistic. By reporting *I^2^*, *τ²* and Cochran’s *Q* statistic we provide a comprehensive assessment of heterogeneity, capturing absolute variance (*τ²*), relative proportion of variability attributable to heterogeneity (*I^2^*), and a formal test of the homogeneity assumption (*Q*).

We conducted an exploratory meta-regression analysis to examine whether study-level variables—such as sample size, mean participant age, exercise intensity, music tempo and the use of self-selected or researcher-selected music—moderated the effect sizes of cognitive control outcomes when combining music with acute exercise, compared to acute exercise alone. A random-effects meta-regression model was fit, with mean participant age and total sample size from each study entered as continuous covariates along with exercise intensity, music tempo, and self-selected vs.researcher-selected music. This approach allowed us to assess the independent influence of these moderators on effect size estimates while accounting for between-study heterogeneity. Data and syntax files for these analyses are available as supplementary files (e.g., OSF https://osf.io/xfn3m/?view_only=ba5de4d156a248998c615f24c17ef001).

## Results

### Study selection

The initial search identified 714 articles. After removing 16 duplicates and four ineligible articles, 645 were excluded based on title and abstract for not meeting inclusion criteria. Forty-seven full articles were assessed, but two were unavailable (as they could not be accessed through institutional subscriptions, with no response received from the authors). Thirty-seven articles were excluded for not using the desired intervention, lacking relevant outcomes, turning out to be secondary reports of other identified studies, and reporting no comparator. In total, 10 articles were deemed eligible for inclusion in the review (Figure 1). Because one study required a minimum of three studies providing data on an outcome to calculate effect sizes, and there was insufficient data for certain identified outcomes, such as working memory (e.g., only two studies reported data for this outcome), nine articles were included in the meta-analysis.

**Figure 1.**
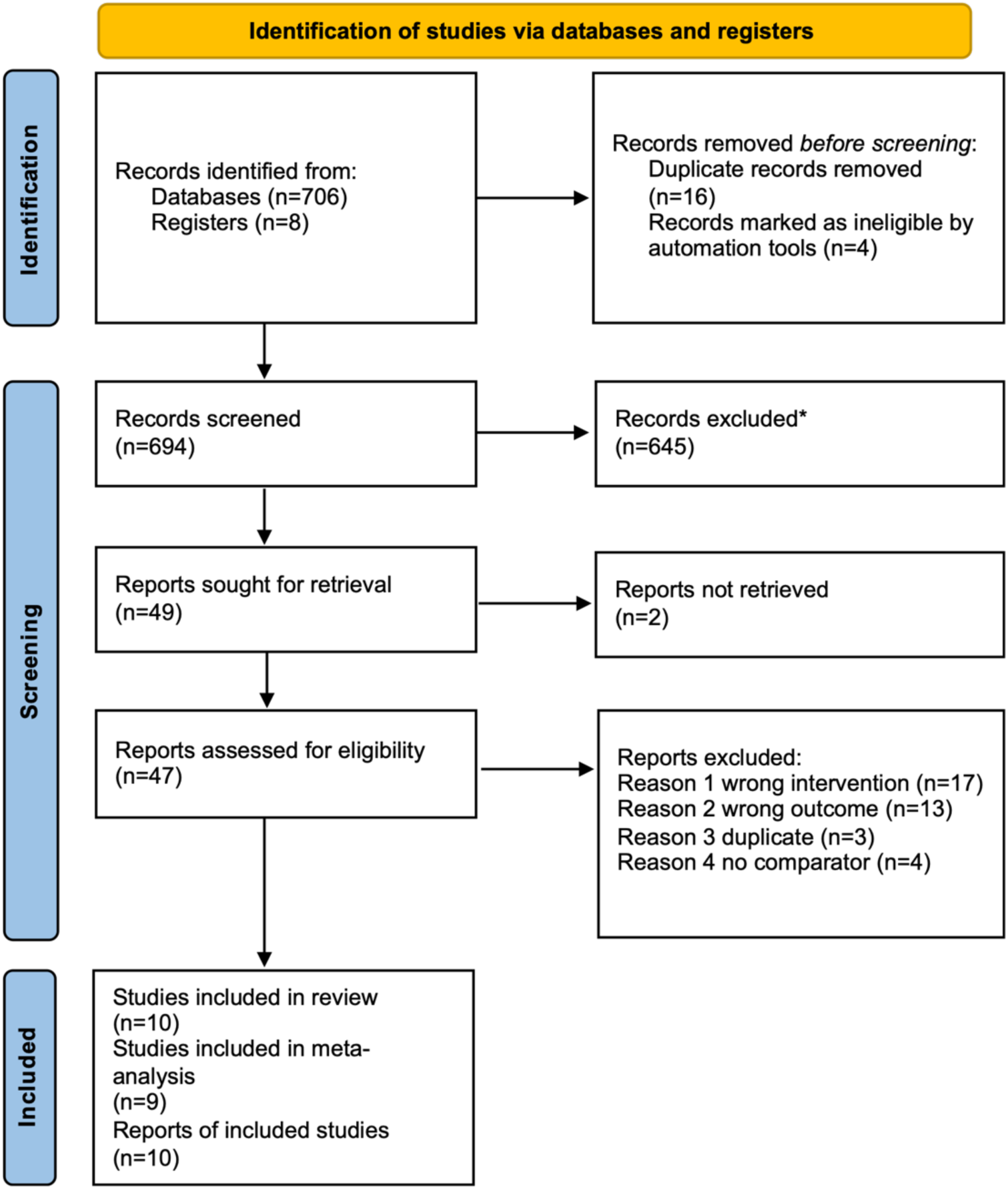
PRISMA information flow describing the screening process. *All records excluded by ASReview [35].

### Study characteristics

The study characteristics (Table 2) include a diverse set of studies conducted in countries such as the United Kingdom (UK) (*n*=2), Brazil (*n*=1), Korea (*n*=1), China (*n*=1), the United States of America (USA) (*n*=2), Japan (*n*=3), and Canada (*n*=1). These studies target a wide range of populations, including healthy young adults, university students, and young adults. A variety of study designs are used, such as RCTs, crossover designs, and repeated measures designs.

**Table 2.**
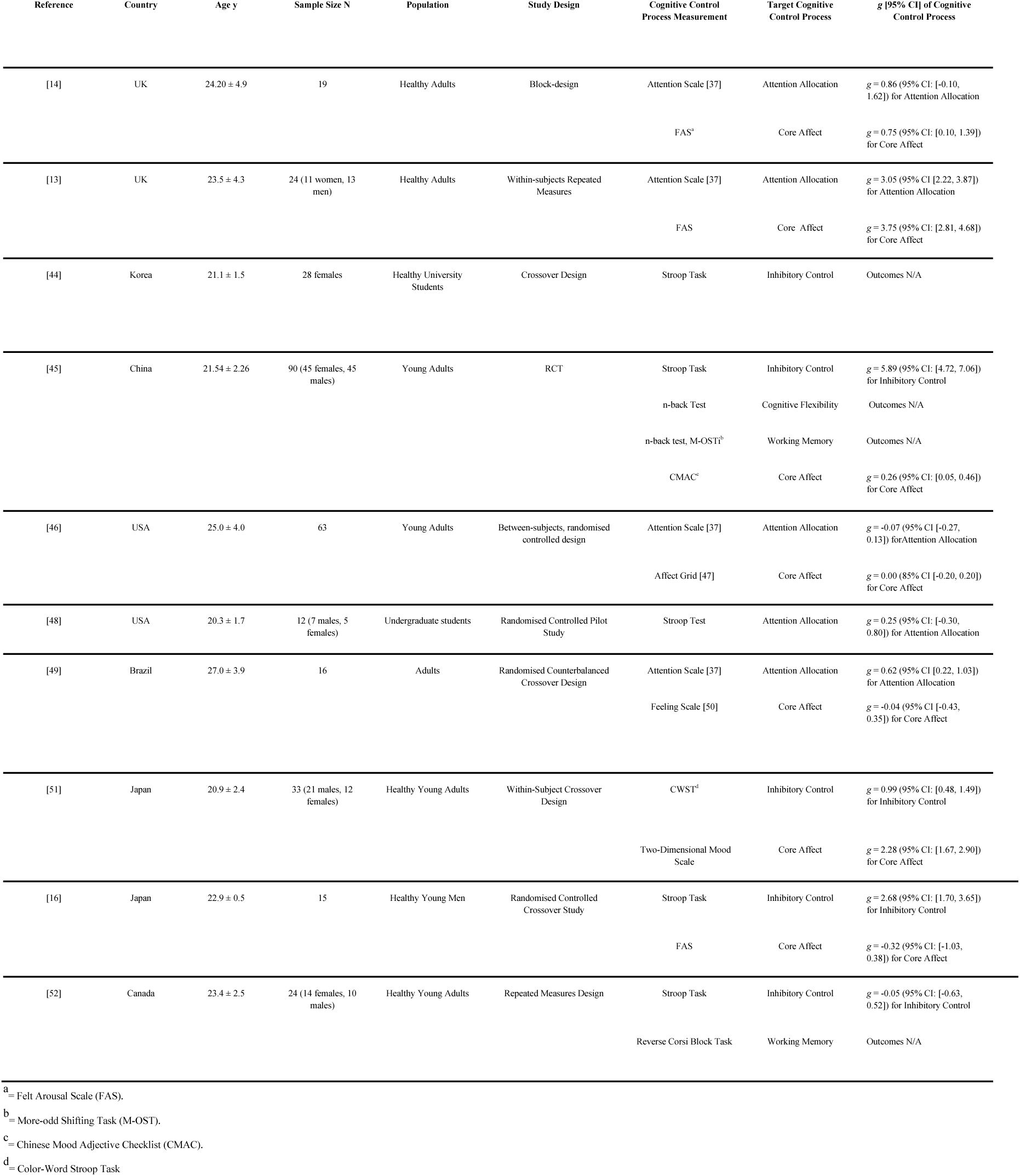
Characteristics of Reviewed Studies.

| Reference | Country | Age $y$ | Sample Size N | Population | Study Design | Cognitive Control Process Measurement | Target Cognitive Control Process | $g$ [95% CI] of Cognitive Control Process |
| --- | --- | --- | --- | --- | --- | --- | --- | --- |
| [14] | UK | 24.20 ± 4.9 | 19 | Healthy Adults | Block-design | Attention Scale [37] | Attention Allocation | $g = 0.86$ (95% CI: [-0.10, 1.62]) for Attention Allocation |
| | | | | | | FAS<sup>a</sup> | Core Affect | $g = 0.75$ (95% CI: [0.10, 1.39]) for Core Affect |
| [13] | UK | 23.5 ± 4.3 | 24 (11 women, 13 men) | Healthy Adults | Within-subjects Repeated Measures | Attention Scale [37] | Attention Allocation | $g = 3.05$ (95% CI [2.22, 3.87]) for Attention Allocation |
| | | | | | | FAS | Core Affect | $g = 3.75$ (95% CI: [2.81, 4.68]) for Core Affect |
| [44] | Korea | 21.1 ± 1.5 | 28 females | Healthy University Students | Crossover Design | Stroop Task | Inhibitory Control | Outcomes N/A |
| [45] | China | 21.54 ± 2.26 | 90 (45 females, 45 males) | Young Adults | RCT | Stroop Task | Inhibitory Control | $g = 5.89$ (95% CI: [4.72, 7.06]) for Inhibitory Control |
|  |  |  |  |  |  | n-back Test | Cognitive Flexibility | Outcomes N/A |
|  |  |  |  |  |  | n-back test, M-OST<sup>b</sup> | Working Memory | Outcomes N/A |
| | | | | | | CMAC<sup>c</sup> | Core Affect | $g = 0.26$ (95% CI: [0.05, 0.46]) for Core Affect |
| [46] | USA | 25.0 ± 4.0 | 63 | Young Adults | Between-subjects, randomised controlled design | Attention Scale [37] | Attention Allocation | $g = -0.07$ (95% CI [-0.27, 0.13]) for Attention Allocation |
| | | | | | | Affect Grid [47] | Core Affect | $g = 0.00$ (85% CI [-0.20, 0.20]) for Core Affect |
| [48] | USA | 20.3 ± 1.7 | 12 (7 males, 5 females) | Undergraduate students | Randomised Controlled Pilot Study | Stroop Test | Attention Allocation | $g = 0.25$ (95% CI: [-0.30, 0.80]) for Attention Allocation |
| [49] | Brazil | 27.0 ± 3.9 | 16 | Adults | Randomised Counterbalanced Crossover Design | Attention Scale [37] | Attention Allocation | $g = 0.62$ (95% CI [0.22, 1.03]) for Attention Allocation |
| | | | | | | Feeling Scale [50] | Core Affect | $g = -0.04$ (95% CI [-0.43, 0.35]) for Core Affect |
| [51] | Japan | 20.9 ± 2.4 | 33 (21 males, 12 females) | Healthy Young Adults | Within-Subject Crossover Design | CWST<sup>d</sup> | Inhibitory Control | $g = 0.99$ (95% CI: [0.48, 1.49]) for Inhibitory Control |
| | | | | | | Two-Dimensional Mood Scale | Core Affect | $g = 2.28$ (95% CI: [1.67, 2.90]) for Core Affect |
| [16] | Japan | 22.9 ± 0.5 | 15 | Healthy Young Men | Randomised Controlled Crossover Study | Stroop Task | Inhibitory Control | $g = 2.68$ (95% CI: [1.70, 3.65]) for Inhibitory Control |
| | | | | | | FAS | Core Affect | $g = -0.32$ (95% CI: [-1.03, 0.38]) for Core Affect |
| [52] | Canada | 23.4 ± 2.5 | 24 (14 females, 10 males) | Healthy Young Adults | Repeated Measures Design | Stroop Task | Inhibitory Control | $g = -0.05$ (95% CI: [-0.63, 0.52]) for Inhibitory Control |
|  |  |  |  |  |  | Reverse Corsi Block Task | Working Memory | Outcomes N/A |

<sup>a</sup> = Felt Arousal Scale (FAS).
<sup>b</sup> = More-odd Shifting Task (M-OST).
<sup>c</sup> = Chinese Mood Adjective Checklist (CMAC).
<sup>d</sup> = Color-Word Stroop Task

### Identification of outcomes and reported measurements of outcomes across studies

The identified outcomes of interest across the studies are: core affect (*n* = 7), inhibitory control (*n* = 6), attention allocation (*n* = 4), working memory (*n* = 2), and cognitive flexibility (*n* = 1) (see Table 3). No outcomes were reported for task switching or overall cognitive performance. Interrater reliability was assessed for the identification of cognitive control process outcomes across the studies, and was found to be acceptable, *k* = 0.667, indicating a moderate to substantial level of agreement among the two raters. Even though initial disagreements were resolved, the moderate reliability demands caution when interpreting the analysis of the identified outcomes of interest.

**Table 3.** Outcomes of interest identified across the included studies.

| Reference | Attention Allocation | Inhibitory Control | Cognitive Flexibility | Working Memory | Core Affect |
| --- | --- | --- | --- | --- | --- |
| [14] | • |  |  |  | • |
| [13] | • |  |  |  | • |
| [44] |  | • |  |  |  |
| [45] |  | • | • | • | • |
| [46] | • |  |  |  | • |
| [48] |  | • |  |  |  |
| [49] | • |  |  |  | • |
| [51] |  | • |  |  | • |
| [16] |  | • |  |  | • |
| [52] |  | • |  | • |  |

We categorised the studies based on specific cognitive processes and measurement tools. Studies under attention allocation, [13,14,46,49] used Tammen’s single-item attention scale [37] to measure associative and dissociative attention. Inhibitory control was assessed using variations of the Stroop task to measure reaction times and error rates in studies by [16,44,45,48,51,52]. Cognitive flexibility was examined using the n-back task by [45].

Similarly, working memory was assessed by [45,52], with [45] using the n-back (1-back and 2-back) and More-odd Shifting Task (M-OST) task, and [52] employing the Reverse Corsi Block Task to examine visuospatial working memory. Core affect was assessed by [13,14,16,45,46,49,51], who used the Feeling Scale (FS) [49], Felt Arousal Scale (FAS) [13,14,16], Affect Grid (AG) [46], Chinese Adjective Mood Checklist (CMAC) [45] and Two-Dimensional Mood Scale (TDMS) [51] to measure affective outcomes. These measures have been commonly used in sports and exercise research domains, with high reliability and validity among them [3]. All studies, with the exception of [16], recorded outcomes from their measurements immediately before or after exercise. [16] measured inhibitory control outcomes before exercise, immediately after exercise, and at three intervals (10, 20, and 30 minutes) during the post-exercise recovery period. Table 4 summarises the measurement tools used across the included studies to assess cognitive control and affective outcomes.

**Table 4.**
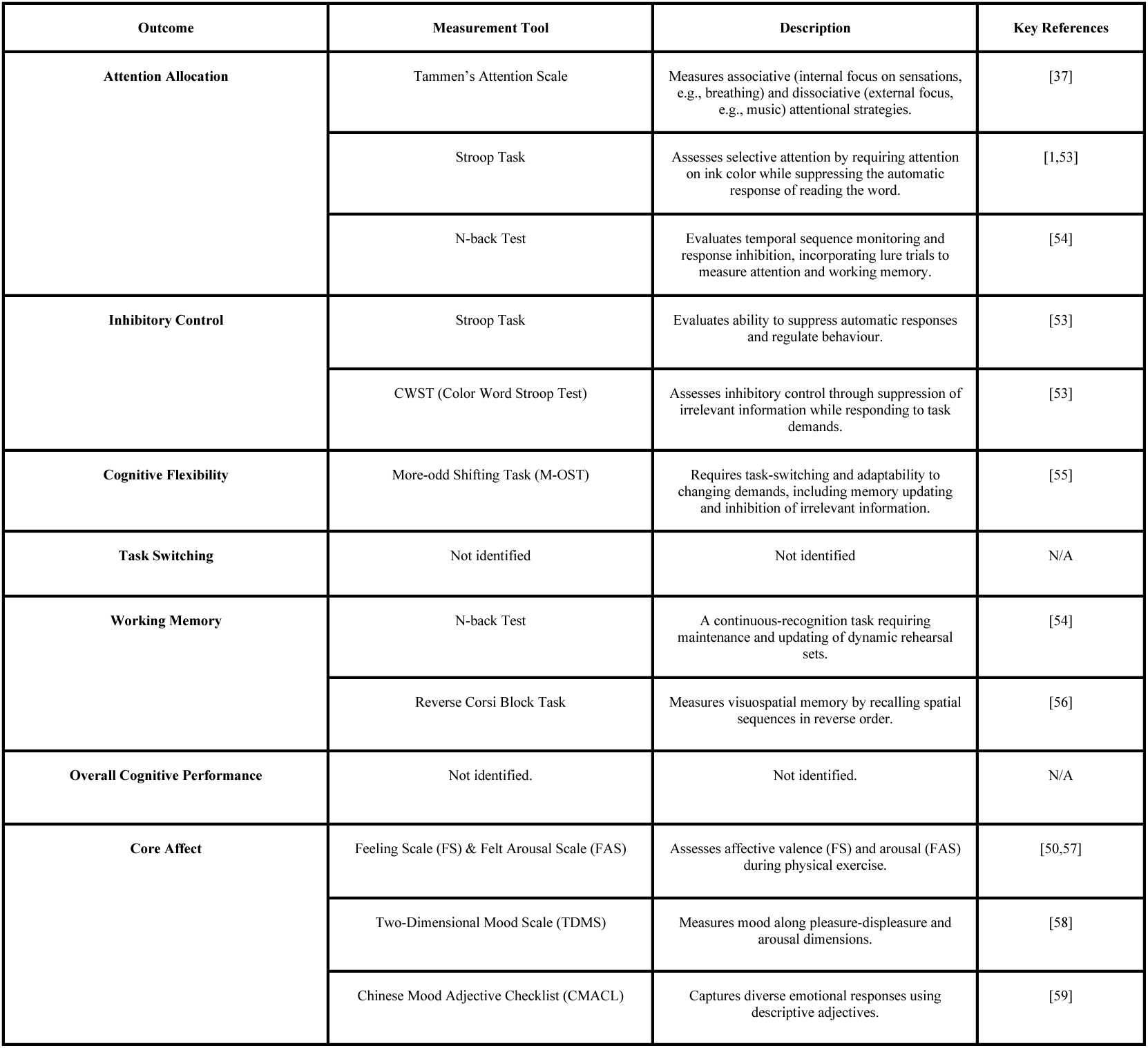
Summary of measurement outcomes for cognitive control and affective outcomes.

| Outcome | Measurement Tool | Description | Key References |
| --- | --- | --- | --- |
| Attention Allocation | Tammen's Attention Scale | Measures associative (internal focus on sensations, e.g., breathing) and dissociative (external focus, e.g., music) attentional strategies. | [37] |
|  | Stroop Task | Assesses selective attention by requiring attention on ink color while suppressing the automatic response of reading the word. | [1,53] |
|  | N-back Test | Evaluates temporal sequence monitoring and response inhibition, incorporating lure trials to measure attention and working memory. | [54] |
| Inhibitory Control | Stroop Task | Evaluates ability to suppress automatic responses and regulate behaviour. | [53] |
|  | CWST (Color Word Stroop Test) | Assesses inhibitory control through suppression of irrelevant information while responding to task demands. | [53] |
| Cognitive Flexibility | More-odd Shifting Task (M-OST) | Requires task-switching and adaptability to changing demands, including memory updating and inhibition of irrelevant information. | [55] |
| Task Switching | Not identified | Not identified | N/A |
| Working Memory | N-back Test | A continuous-recognition task requiring maintenance and updating of dynamic rehearsal sets. | [54] |
|  | Reverse Corsi Block Task | Measures visuospatial memory by recalling spatial sequences in reverse order. | [56] |
| Overall Cognitive Performance | Not identified. | Not identified. | N/A |
| Core Affect | Feeling Scale (FS) & Felt Arousal Scale (FAS) | Assesses affective valence (FS) and arousal (FAS) during physical exercise. | [50,57] |
|  | Two-Dimensional Mood Scale (TDMS) | Measures mood along pleasure-displeasure and arousal dimensions. | [58] |
|  | Chinese Mood Adjective Checklist (CMACL) | Captures diverse emotional responses using descriptive adjectives. | [59] |

Each tool is linked to specific constructs with key references.

### Study categorisation into meta-analysis clusters

As the Stroop task is not a pure measure of either inhibitory control or attention allocation [43], we differentiated between studies focusing on inhibitory control and those examining attention allocation, specifically organising them into two distinct meta-analysis clusters. For attention allocation, studies were required to use a variation of the Stroop task as a measure of selective attention, emphasising reaction times, particularly the Stroop effect (i.e., the difference between incongruent and congruent trial times), as the primary dependent variable. These studies needed to explicitly state their aim of investigating attention allocation or selective attention in the Stroop context. For inhibitory control, studies also had to employ the Stroop task but emphasise Stroop interference scores (which quantify the suppression of participants’ automatic responses). These studies had to clearly indicate a focus on inhibitory control or prepotent response suppression within the Stroop paradigm.

Inclusion to this cluster was extended to studies incorporating other inhibitory control tasks, such as the Flanker or Go/No-Go tasks. Core Affect was also included as a separate cluster. Following the Circumplex Model of Affect [29], we categorised affective responses engendered by music along two dimensions: valence, which represents the spectrum of pleasure to displeasure, and arousal, which entails the level of activation or energy.

### Risk of Bias in Studies

Following the assessment of the study quality using the JBI critical appraisal checklist tools [36], the nine criteria were adapted to the five risk of bias domains found in the [52] R package for risk-of-bias assessments (robvis). This assessment tool evaluates the risk of bias resulting from the randomisation process (D1), deviations from intended intervention (D2), missing outcome data (D3), measurement of the outcome (D4), and selection of the reported result (D5). Each domain is assessed using a judgement scale indicating high risk of bias (red cross), some concerns (yellow circle), low risk of bias (green plus), and No Information (blue question mark), with a summary of the overall risk of bias for each study presented in Figure 2. The overall risk of bias rating was determined by assigning the lowest rating observed across any of the five domains (i.e., if a high risk was present in any domain, the overall rating was also high).

**Figure 2.**
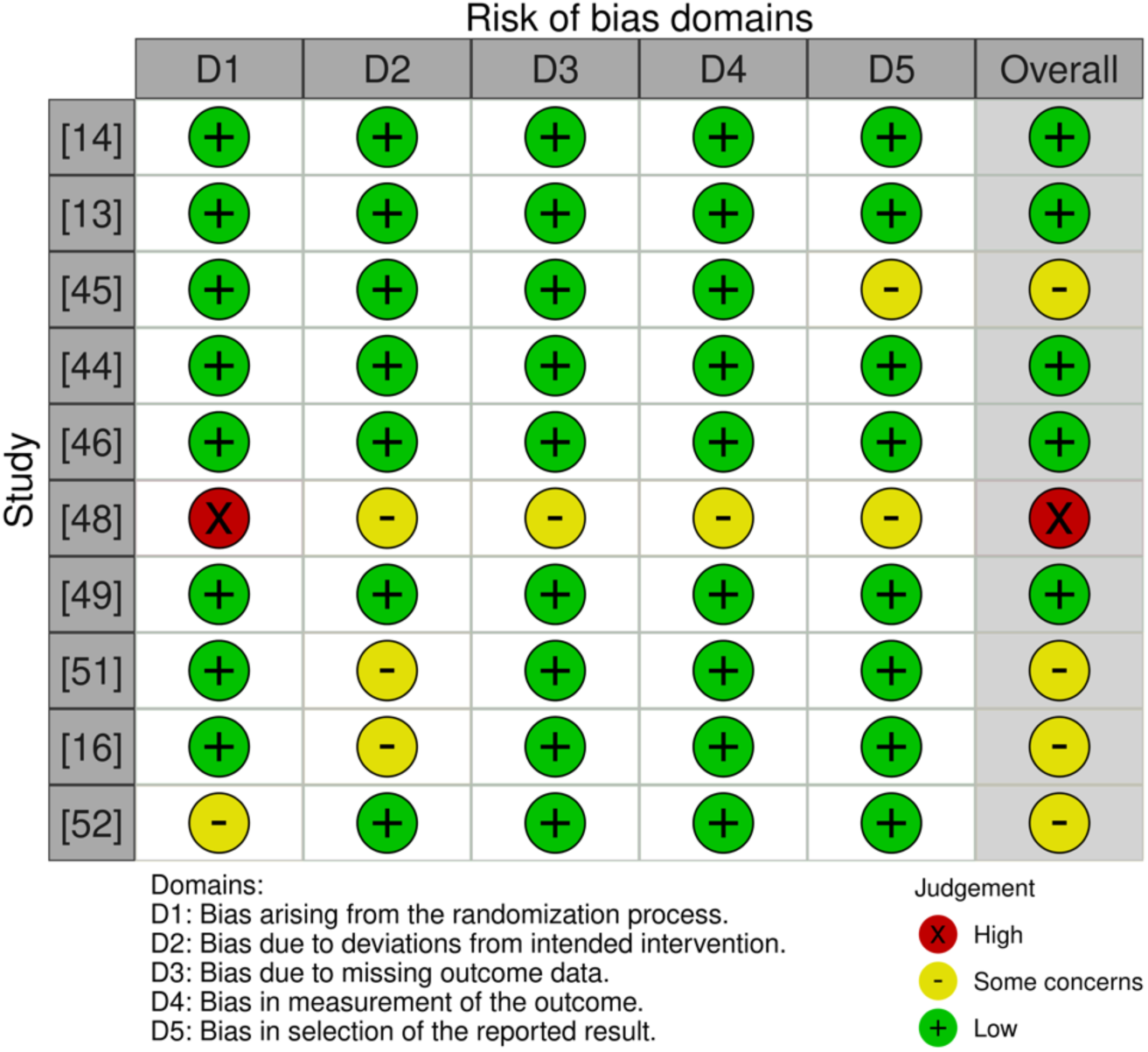
Evaluation of risk of bias in the studies included in the review, categorised across five domains from D1 to D5. An overall bias risk assessment for each study is also provided, summarising the findings across all domains. [60].

Of the 10 studies, five studies were rated for a low risk of bias. Four studies received a moderate (some concerns) rating, and one study received a high rating in risk of bias. We included all studies in the review regardless of their quality rating (Figures 2 and 3).

**Figure 3.**
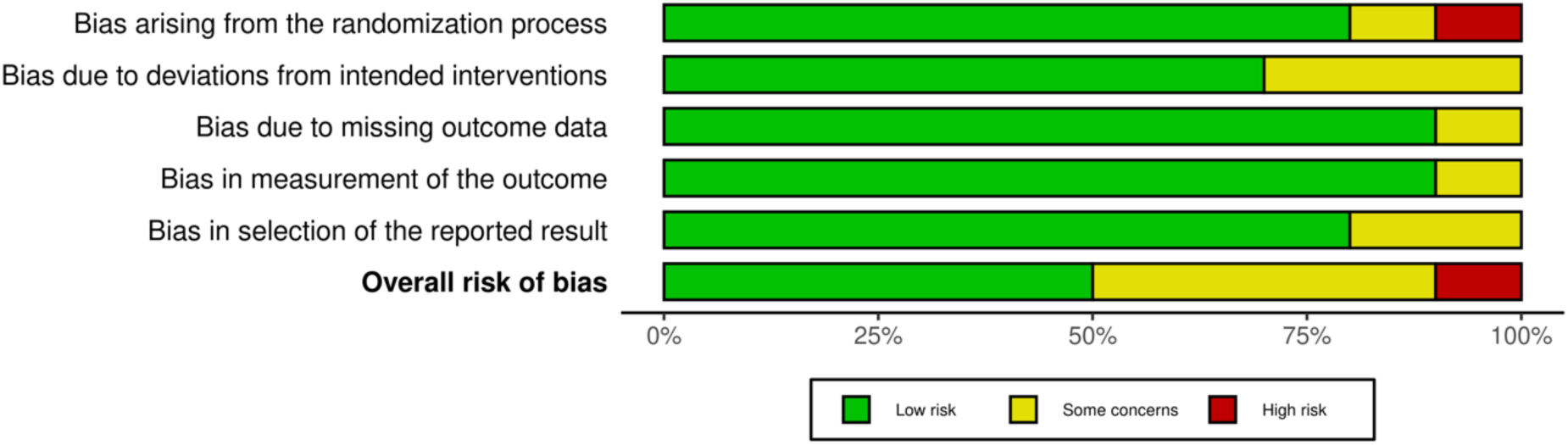
The overall risk of bias for each study is also summarised, with green representing low risk, yellow indicating some concerns and red representing high risk. The majority of studies fall within low risk for most domains, though some domains exhibit higher proportions of “some concerns” for bias. [60].

### Narrative synthesis of cognitive control processes

Individual study findings were analysed qualitatively across the following identified outcomes of interest, attention allocation, inhibitory control, cognitive flexibility, working memory and core affect.

### Attention allocation

Music consistently activated dissociative attention during low- to moderate-intensity exercise and recovery periods, reducing the salience of interoceptive cues such as fatigue.

However, the effectiveness of music in shifting attention externally varied depending on study protocols and exercise modalities. For instance, [14] examined 19 young adults performing submaximal isometric handgrip contractions in a functional magnetic resonance imaging (fMRI) scanner and found increased neural activation in sensory and attentional control regions (e.g., the left inferior frontal gyrus) concurrent with music listening (i.e., music was played during the fMRI scan). In contrast, [13] studied a broader sample of physically active adults cycling at moderate intensity and found that music promoted subjective dissociative attention, but this was reported through self-reported measures rather than neuroimaging. Similarly, [46] found that both fast- and slow-tempo music supported dissociative attention during a wall-sit exercise but not during a plank hold, indicating that the effectiveness of music varies across isometric tasks. During sprint interval training (SIT), [49] reported that music enhanced dissociative attention in recovery periods but had no significant effect during bouts of high intensity exercise, indicating music’s conditional utility (e.g., music may aid recuperation and recovery during low-intensity exercise phases, but is less effective in diverting attention allocation during high exercise intensities). Collectively, the evidence suggests that music facilitates dissociative attention in moderate-intensity dynamic tasks (e.g., cycling) or recovery phases (low intensity exercise), but its effects diminish in static (e.g., plank hold exercise) or high-intensity conditions, where associative attention may dominate.

### Inhibitory control

Mixed results on inhibitory control were found as music was combined with acute exercise, with outcomes influenced by exercise intensity, protocol design, and the presence of auditory-motor coupling. [45] showed that tempo-matched music synchronised to participants’ heart rates during moderate-intensity aerobic exercise, significantly influenced inhibitory control, likely due to optimised auditory-motor coupling (e.g., participants exhibited improved Stroop task performance when cycling at a tempo aligned with music at 120–140 BPM, facilitating synchronisation between their heart rate and the auditory rhythm). Similarly, [44] observed differential effects of high-decibel music during high-intensity exercise in a smaller sample of resistance-trained females. However, null results were reported in [48,51]: [51] found no significant differences between music and metronome conditions on inhibitory control following moderate-intensity cycling, while [48] observed no such effects of music or high-intensity interval training on inhibitory control, suggesting that task sensitivity and protocol alignment (e.g., matching music tempo to exercise intensity) may be most salient to influence this outcome. Variability in participant characteristics further contributed to inconsistencies, with [45] recruiting a larger, more diverse sample compared to smaller, demographically specific cohorts in [44,48]. The mixed evidence indicates that music’s effects on inhibitory control depend on contextual factors such as task design, exercise intensity, and the presence of auditory-motor coupling.

### Cognitive flexibility

One RCT [45] examined cognitive flexibility using a combined music (i.e., pop music without lyrics, with three different tempi, 60-65 BPM, 120-140 BPM and 155-165 BPM) and exercise protocol (i.e., a 20-minute bout of moderate-intensity aerobic exercise on a bicycle ergometer at 50–60 revolutions per minute, RPM) with 90 young adults. Although no strict no-music condition was included, the slower mismatched (60-65 BPM) and faster mismatched tempo (155-165 BPM) groups could serve as proxies for a no-music condition (as indicated in our eligibility criteria). Results indicated no significant differences in cognitive flexibility outcomes across groups, suggesting that tempo variations did not critically influence this outcome.

### Working memory

[45] found that music matched in tempo (120-140 BPM) with the exercise routine significantly improved working memory outcomes in 30 young adults, compared to slower and faster music tempi. In contrast, [52] found no significant improvements in working memory between exercise (a 30-minute session on a recumbent cycle ergometer at a moderate intensity, 55% of the participant’s heart rate reserve) alone and exercise with music (a musical playlist consisting of classical songs with a tempo ranging from 120-140 BPM) in 24 young adults. The effects of music and acute exercise on working memory are insufficiently differentiated across conditions to draw definitive conclusions. Some evidence suggests that factors such as alignment between music tempo and exercise rhythm, the type of exercise performed, and individual participant characteristics may determine whether working memory outcomes improves or remains unaffected.

### Core affect

[14] demonstrated that music significantly elevated affective arousal by engaging the left inferior frontal gyrus, fostering a dissociative state, reducing perceived exertion, and producing more pleasurable participant experience during light-to-moderate intensity exercise. Similarly, [45] report that tempo-matched music – a form of auditory-motor coupling – elevated affective valence and during moderate-intensity aerobic exercise, where synchronised music tempo and participant physiological rhythms (e.g., heart rate) definitively improved the effects of music on core affect outcomes, compared to slower or faster-matched music.

Exercise intensity emerged as a critical factor influencing these outcomes. [46,49] reported no significant changes in affective valence or arousal during high-intensity or isometric exercises, likely due to the dominance of physiological stressors (e.g., fatigue, physical exertion), which potentially diminish music’s effectiveness under high physical demand.

Concurrent with exercise intensity, exercise protocols influence outcomes: while tempo-matched music was particularly effective during sustained moderate-intensity efforts [14,45], its effects were less pronounced during short, high-intensity sprints or isometric tasks [46,49], where those physical demands likely outweighed music’s differential cognitive and affective effects. Overall, the affective responses to music and acute exercise evident in the literature reviewed indicate a context-dependent influence on core affect outcomes, elevating affective arousal at moderate intensities (e.g., tempo-matched music during steady-state aerobic exercise) but yielding inconsistent effects during high-intensity or physiologically demanding exercise protocols (e.g., sprint interval training or isometric tasks).

### Meta-analysis

A single overall meta-analysis was not conducted due to substantial heterogeneity across datasets and outcomes. Instead, outcomes were categorised into distinct domains for separate analyses: (1) attention allocation (*n* = 4), (2) inhibitory control (*n* = 5), and (3) core affect (*n* = 7). Meta-analyses could not be conducted on cognitive flexibility (*n* = 1) and working memory (*n* = 2) due to the limited number of studies and data reported for each process.

### Results for attention allocation

The overall effect size was 1.05, with a 95% confidence interval (CI) of 0.03 to 2.07 and a *p*-value of 0.043 (*k* = 4, *n* = 122) (Figure 4). These results were statistically significant, supporting music during acute exercise to have a substantial effect on attention allocation outcomes. The random-effects model revealed high heterogeneity between studies, indicating substantial variability across the included studies (*Q* = 59.732, *p* < 0.001, *I²* = 94.98%, *Tau²* = 0.992). Furthermore, the 95% prediction interval ranged from -5.13 to 7.23, indicating the wide variability and uncertainty in the true effects that could be observed in future studies.

**Figure 4.**
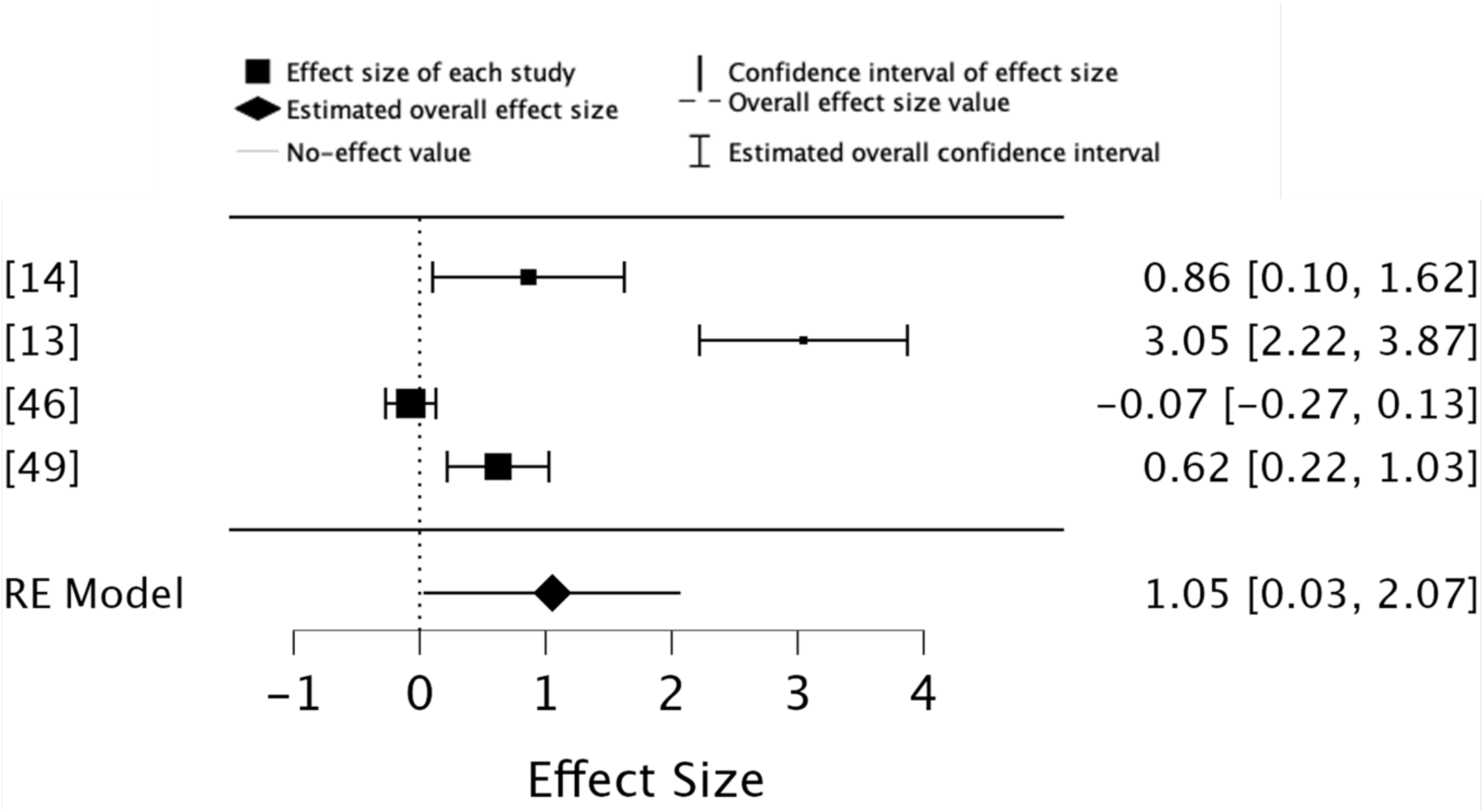
Forest plot of effect sizes for attention allocation outcomes of music during acute exercise compared to no music during acute exercise.

### Results for inhibitory control

The overall effect size was 1.87, with a CI of 0.37 to 3.37 and a *p*-value of 0.014 (*k* = 5, *n* = 174), indicating that the results were statistically significant and supported the notion that music has a substantial effect on inhibitory control outcomes during exercise (Figure 5). The random-effects model revealed substantial heterogeneity (*Q* = 99.063, *p* < 0.001, *I²* = 95.96%, *Tau²* = 2.757), suggesting a high degree of variability across the studies. The 95% prediction interval ranged from -1.32 to 5.06, indicating a broad variability in the expected true effect sizes for future studies.

**Figure 5.**
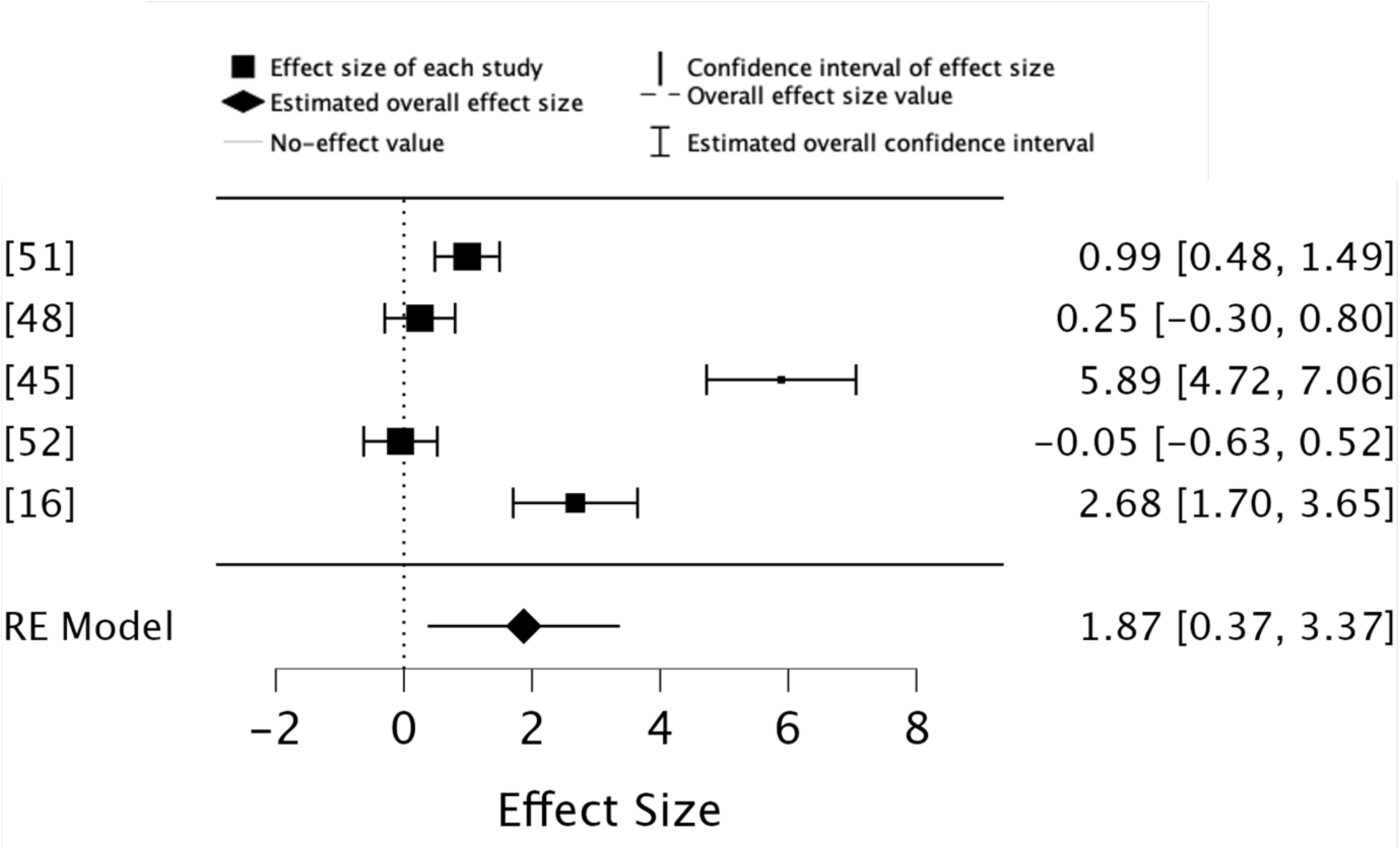
Forest plot illustrating the effect sizes of studies comparing inhibitory control outcomes of music during acute exercise compared to no music during acute exercise.

A sensitivity analysis, which excluded an outlier study with a large effect size ([45] *g* = 5.89), produced an overall effect size of 0.89, with a CI of -0.02 to 1.80 and a *p*-value of 0.054 (*k* = 4, *n* = 144). This result was not statistically significant, indicating that [45] may have had a disproportionate influence on the overall findings regarding inhibitory control outcomes during acute exercise (Figure 6). The random-effects model indicated reduced but still substantial heterogeneity (*Q* = 26.241, *p* < 0.001, *I²* = 88.57%, *Tau²* = 0.748). The 95% prediction interval ranged from -3.42 to 5.20, highlighting the wide variability in the expected true effect sizes for future studies.

**Figure 6.**
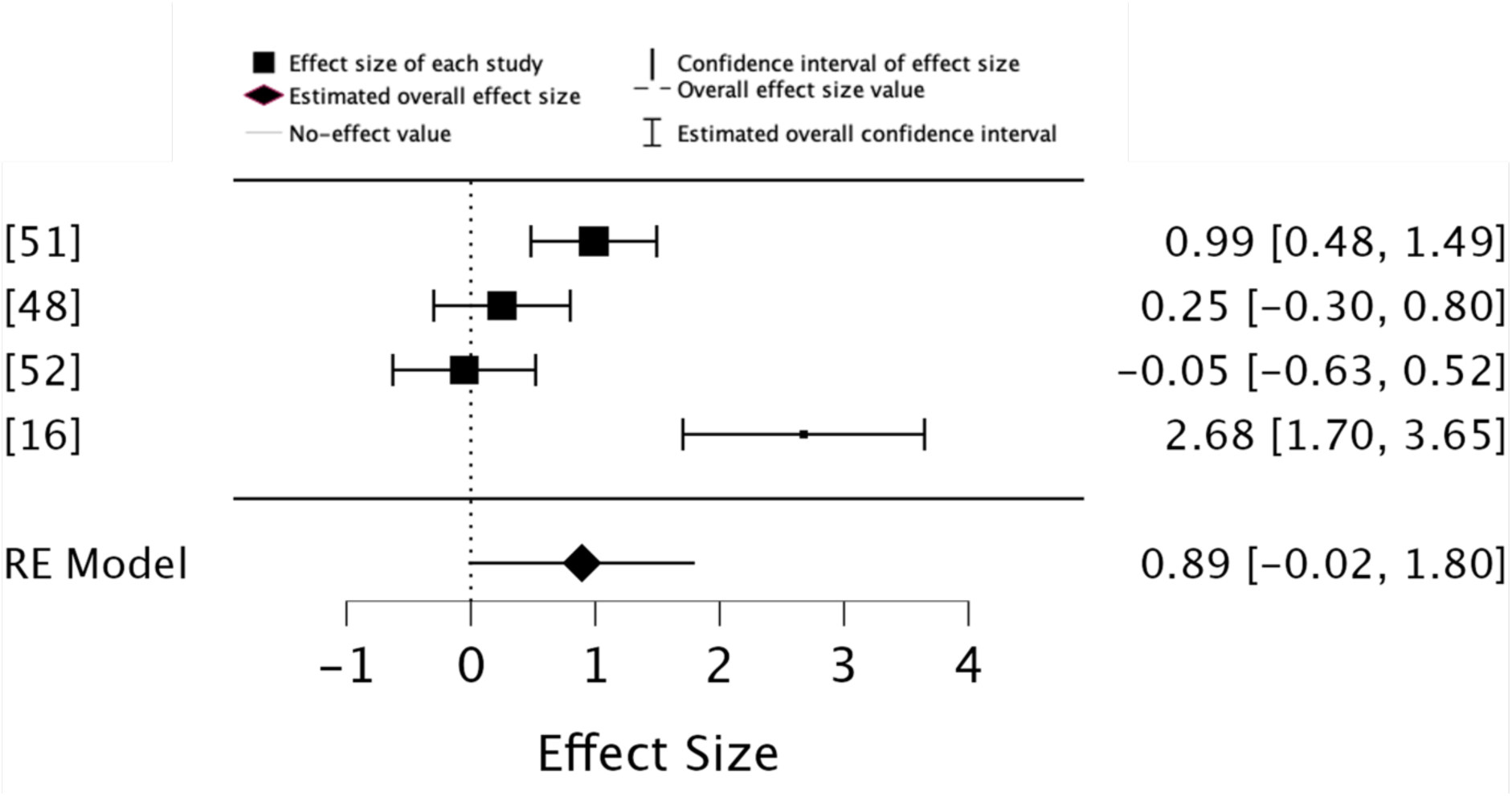
Forest plot of a sensitivity analysis excluding [45], illustrating the effect sizes of studies comparing inhibitory control outcomes with music during acute exercise to no music during acute exercise.

### Results for core affect

The overall effect size was 0.86, with a CI of 0.24 to 1.48 and a *p*-value of 0.007 (*k* = 7, *n* = 200), indicating that the results were statistically significant and supported the notion that music has a significant effect on affect outcomes during exercise (Figure 7). The random-effects model indicated high heterogeneity across studies (*Q* = 108.724, *p* < 0.001, *I²* = 94.48%, *Tau²* = 0.618), suggesting considerable variability between the studies. Additionally, the 95% prediction interval ranged from -1.38 to 3.10, highlighting that while the average effect size is positive, future studies may observe a wide range of effects.

**Figure 7.**
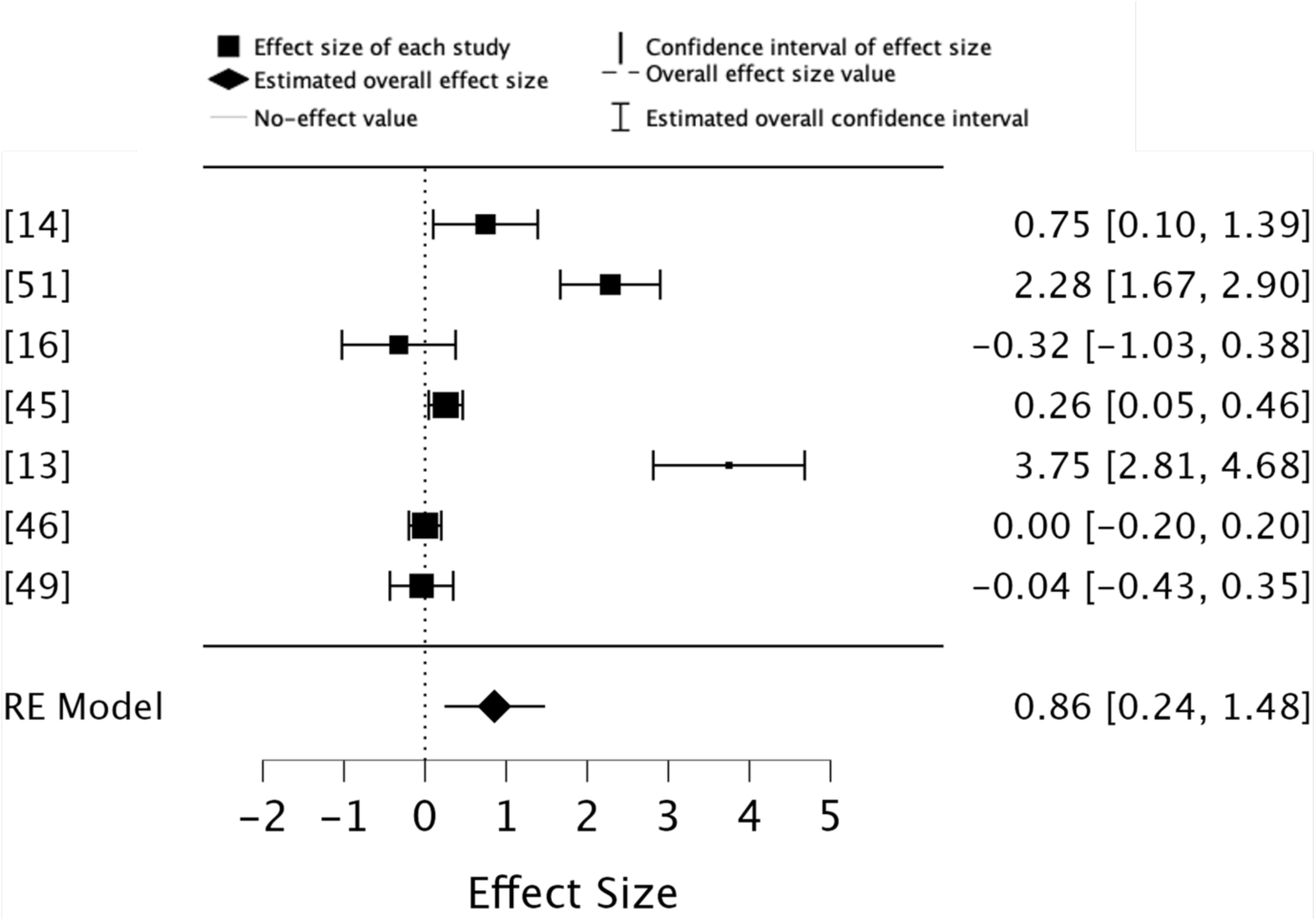
Forest plot showing the effect sizes of studies comparing core affect outcomes of music during acute exercise compared to no music (and proxies) during acute exercise.

A sensitivity analysis, which excluded one study by [13] due to its large effect size (*g* = 3.75), produced an overall effect size of 0.45, with a CI of -0.04 to 0.94 and a *p*-value of 0.069 (*k* = 6, *n* = 176) (Figure 8). This result was not statistically significant, indicating that the outcomes of [13] may have substantially influenced the overall core affect results. The random-effects model indicated high residual heterogeneity (*Q* = 54.476, *p* < 0.001, *I²* = 90.82%, *Tau²* = 0.308), suggesting considerable variability between the studies even after removing the outlier. The 95% prediction interval ranged from -0.093 to 0.993, indicating that the true effect sizes in future studies may vary widely.

**Figure 8.**
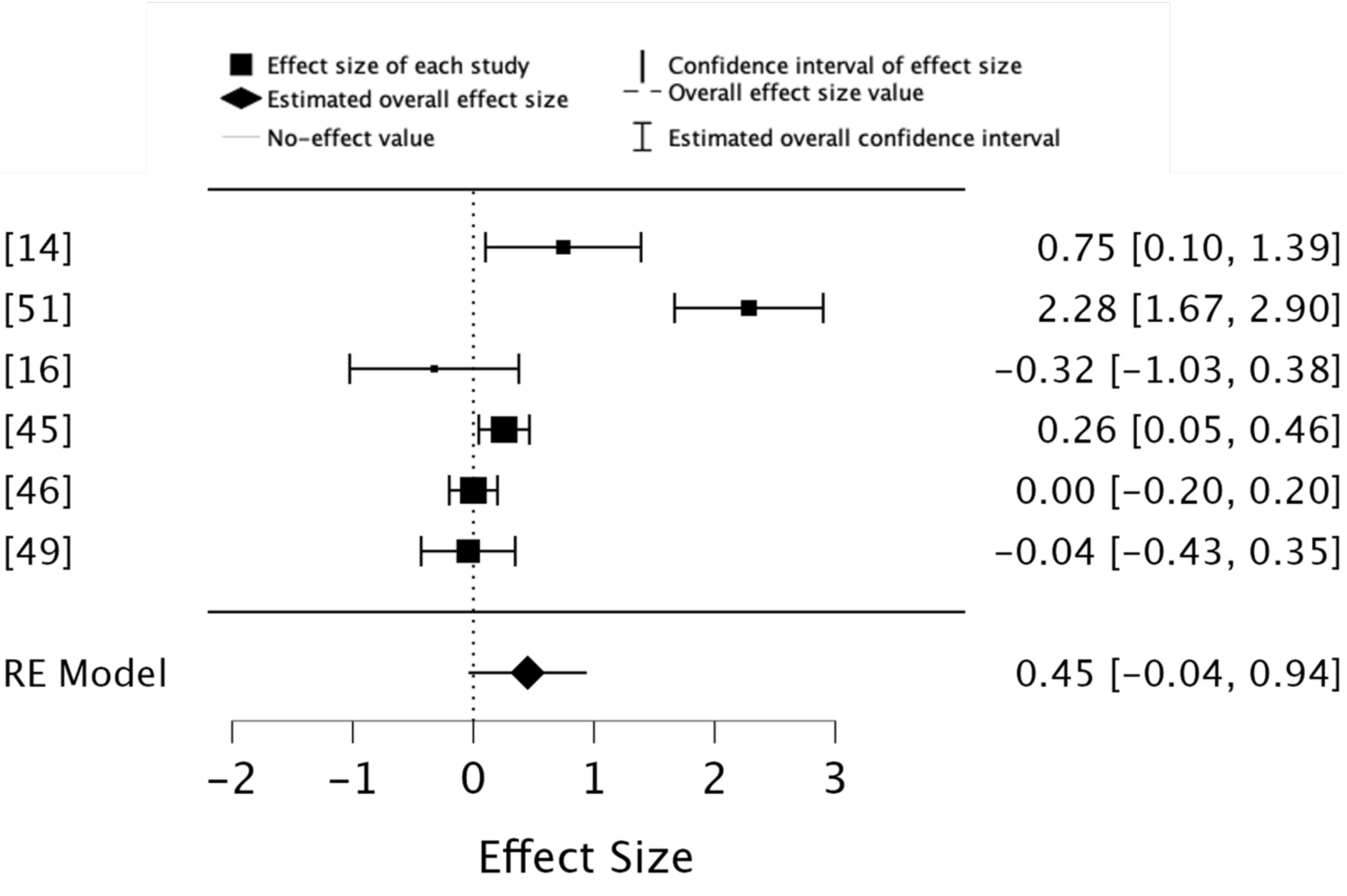
Forest plot of a sensitivity analysis excluding [13], illustrating the effect sizes of studies comparing core affect outcomes with music during acute exercise to no music (or proxies) during acute exercise.

### Sub group analysis results: affective arousal

The overall effect size was 1.37, with a CI of -0.74 to 3.74 and a *p*-value of 0.204 (*k* = 3, *n* = 58) (Figure 9). This result was not statistically significant, suggesting insufficient evidence to support a significant effect of music used during acute exercise on affective arousal outcomes. The random-effects model indicated substantial heterogeneity (*Q* = 47.558, *p* < 0.001, *I²* = 95.80%, *Tau²* = 3.317), highlighting significant variability across studies. The 95% prediction interval ranged from -5.81 to 8.55, indicating a wide range of potential true effect sizes in future studies.

**Figure 9.**
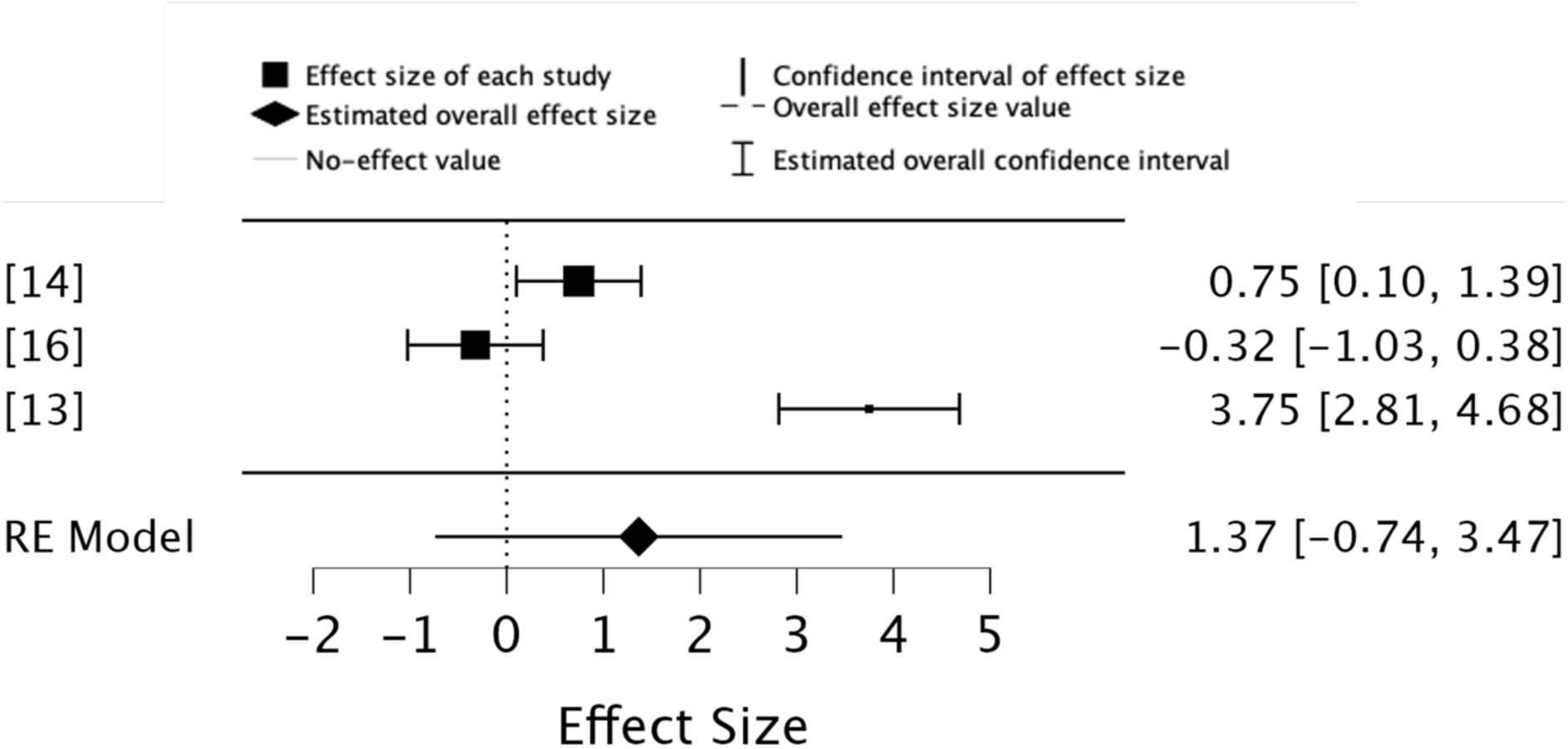
Forest plot showing the effect sizes of studies comparing affective arousal outcomes of music during acute exercise compared to no music (and proxies) during acute exercise.

### Sub group analysis results: affective valence

The overall effect size was 0.79, with a CI of -0.22 to 1.80 and a *p*-value of 0.124 (*k* = 3, *n* = 79) (Figure 10). This result was not statistically significant, suggesting insufficient evidence to support a significant effect of music used during acute exercise on affective valence outcomes. The random-effects model indicated substantial heterogeneity (*Q* = 42.557, *p* < 0.001, *I²* = 95.30%, *Tau²* = 0.744), highlighting significant variability across studies. Furthermore, the 95% prediction interval ranged from -6.39 to 7.97, indicating a wide range of potential true effect sizes in future studies.

**Figure 10.**
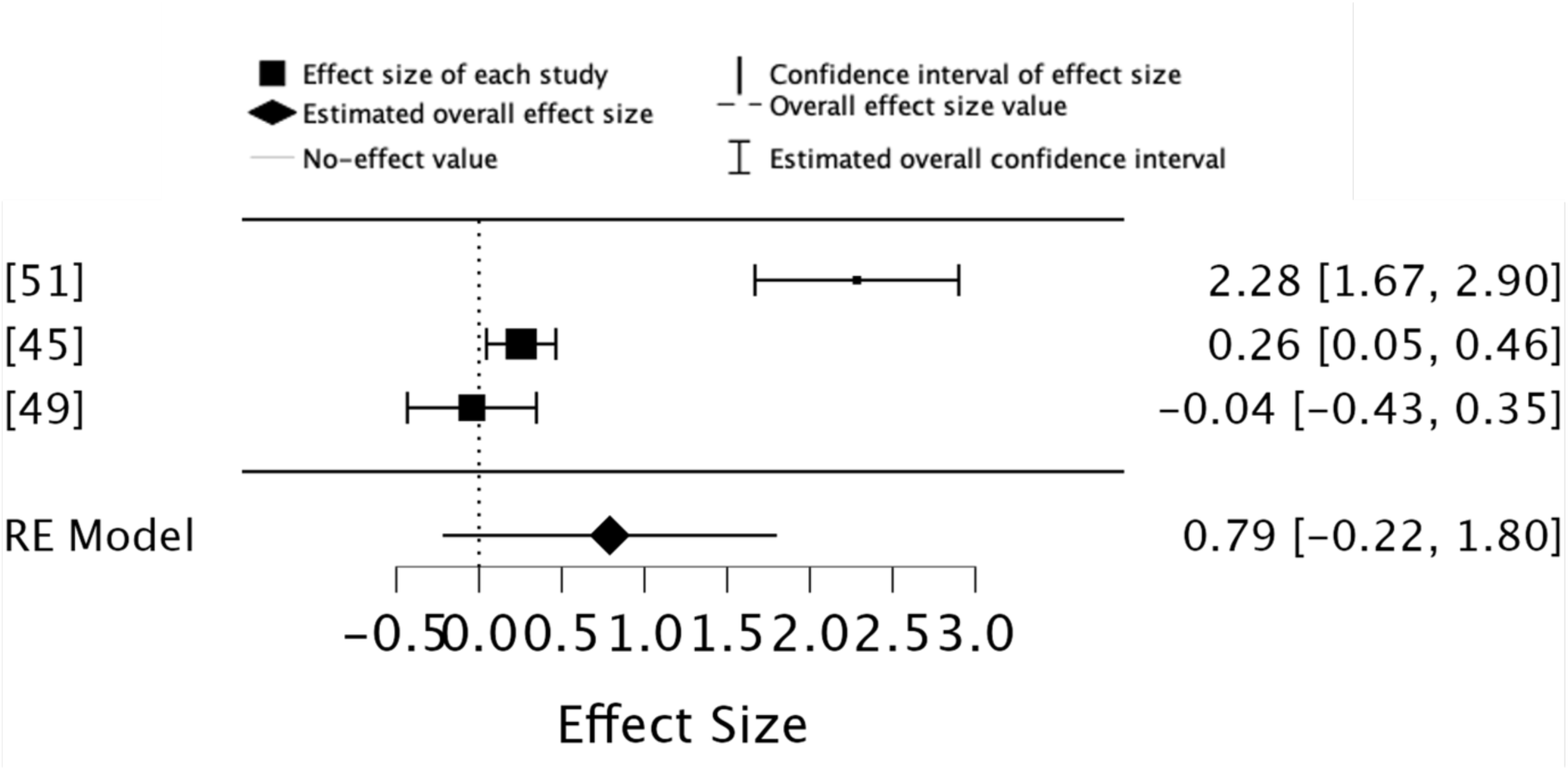
Forest plot showing the effect sizes of studies comparing affective valence outcomes of music during acute exercise compared to no music (and proxies) during acute exercise.

### Meta-regression analysis

Heterogeneity was identified in all meta-analyses. Therefore, a meta-regression analysis was performed to explore study size, mean participant age, exercise intensity, music tempo, and the use of self-selected or researcher-selected music (music selection) as potential moderators of effect sizes, comparing combined music and exercise protocols with exercise alone. In our meta-regression analysis, exercise intensity, music tempo and the use of self-selected music or researcher-selected music (music selection) were coded as categorical variables to facilitate comparison across studies. For exercise intensity, studies reporting intensity in broad categories such as low (e.g., walking at 3–4 km/h or 30–40% VO2 max), moderate (e.g., jogging at 5–6 km/h or 40–60% VO2 max), or high (e.g., running at 7–8 km/h or above 70% VO2 max) [61] were assigned numeric values (1 = Low, 2 = Moderate, 3 = High). Music tempo was categorised based on beats per minute (BPM) into three distinct groups (1 = Slow [60–90 BPM]; 2 = Medium [91–130 BPM]; 3 = Fast [131+ BPM]). The use of self-selected music and researcher-selected music was coded in two groups (1 = Self-selected music; 2 = Researcher-selected music). The analysis encompassed all outcomes of interest due to the small number of studies available for meta-analyses.

Exercise intensity yielded a significant effect on the effect sizes (*p* = .024), with higher exercise intensities reducing the effectiveness of music in enhancing cognitive control outcomes. None of the other variables had a statistically significant impact on the effect sizes (see Table 5). The overall model remained non-significant (*p* = .147), and substantial heterogeneity persisted (*Q* = 150.095, *p* < 0.001, *I²* = 98.71%, *Tau²* = 2.055). This suggests that unexplored factors may contribute to the heterogeneity in outcomes. Findings should be interpreted cautiously, as the analysis included all outcomes of interest, increasing the potential for variability that was not accounted for.

**Table 5.** A summary of the meta-regression analysis.

|  | Estimate | Standard Error | *z* | *p* |
| --- | --- | --- | --- | --- |
| Intercept | 7.153 | 4.960 | 1.442 | .149 |
| Study Size | -0.029 | 0.027 | -1.082 | .279 |
| Participant Age | -0.053 | 0.037 | -1.441 | .150 |
| Exercise Intensity | -1.768 | 0.784 | -2.255 | .024 |
| Music Tempo | 0.303 | 0.948 | 0.320 | .749 |
| Music Selection | -0.512 | 1.018 | 0.502 | .615 |

*Note.* Wald test.

## Discussion

This review systematically evaluated the efficacy of combining music with acute exercise by exploring its potential effect on cognitive control processes and affective outcomes. Exploratory meta-analyses show significant effects on attention allocation, inhibitory control and core affect outcomes, consistent with prior meta-analyses on the cognitive [5] and affective effects of combined music and exercise [3]. Meta-regression analysis identified exercise intensity as a significant moderator, with higher exercise intensities attenuating the positive effects of music on cognitive and affective outcomes. This concurs with evidence [20,24] that low-to-moderate exercise intensities optimise differential effects on cognitive processes, while higher intensities may impose excessive physiological and cognitive demands, potentially diminishing such effects [20,24].

The overall certainty of this evidence is limited by methodological inconsistencies, a small number of eligible studies, and the substantial heterogeneity observed across studies. Sensitivity analyses highlight the disproportionate influence of individual studies, such as study [45] in the inhibitory control cluster, which significantly influenced overall effect sizes. Due to the aforementioned heterogeneity, the findings of this review must be interpreted cautiously. While prediction intervals reveal substantial variability in potential true effect sizes—ranging from negative to large positive effects—this variability reflects the nuanced, context-dependent relationship between music, exercise, cognitive control processes and affective responses. The heterogeneity indicates the complexity of these interactions, as well as the potential for individualised responses, warranting further investigation. Despite variability, positive trends in specific cognitive control processes such as attention allocation, inhibitory control and affective outcomes, highlight the potential value of the music-exercise combination, pointing to the need for high-quality research to refine these findings.

### Potential mechanisms of music-exercise effects on cognitive control and affective outcomes

This study suggests that the intensity of acute exercise moderates the influence of music on cognitive control and affective outcomes. Specifically, low-to-moderate exercise intensities provide favourable conditions for music to enhance cognitive control processes and improve affective responses. This intensity-dependent influence likely stems from several factors. At low-to-moderate exercise intensities, the cognitive demands of exercise are lower, leaving more cognitive resources available for processing external stimuli such as music [38]. This enables individuals to better engage with music, as it redirects their attentional resources, reduces interoceptive cueing (e.g., physical exertion), and fosters elevated affective responses [50]. Music’s ability to drive and synchronise physiological rhythms (e.g., through auditory-motor coupling, where auditory stimuli influence motor responses) further enhances cognitive outcomes, as demonstrated by improved Stroop task performance in studies where participants’ heart rates aligned with a tempo set by the music [8,45]. These effects extend to affective outcomes, where tempo-matched music with acute exercise was shown to elevate core affective responses, improving the pleasantness of the task [45].

The observed cognitive and affective benefits of music during low-to-moderate exercise intensity align with DMT’s assertion that low-to-moderate exercise intensities allow for consistent cognitive-affective interaction within exercise, due to tolerable physiological and cognitive demands [19,23,26]. An interesting and somewhat unexpected finding to emerge from the meta-analysis data were the null results for the sub group analysis of affective valence (see Figure 10), potentially attributable to the high interoceptive demands of exercise in study [49], mismatches in music tempo [45] or music selection [16], as well as variability in participant preferences and measurement methods across the included studies.

This hints that the affective arousal dimension plays a more critical role in influencing cognitive processes (e.g., attention allocation, inhibitory control) and affective outcomes, compared to affective valence – particularly at low-to-moderate exercise intensities where arousal is optimally modulated by music without overwhelming cognitive resources [13,14,26]. The null affective arousal results (see Figure 9) may reflect participants already at optimal arousal levels (as these studies utilised low-to-moderate exercise intensity protocols [13,14,16]), leaving little room for music to influence it. Taken together, these findings provide insight into how the cognitive and affective domains of music-exercise are constrained by intensity-dependent physiological and interoceptive demands that surpass the zone of response variability [26].

The narrative synthesis points to the contextual nature of these effects. For instance, [44] reported benefits of high-decibel music during high-intensity exercise, while [48] observed no significant effects of music in similar conditions, suggesting that mismatches in exercise intensity, music tempo, and task demands may negate music and acute exercise potential effects. Similarly, [51] found that auditory stimuli such as metronomes were as effective as music, indicating that the type of auditory stimulus may play a lesser role compared to its synchronisation with motor activity. Regarding affective outcomes, mismatches between music tempo and participant musical preferences [16] or the use of high-intensity protocols [46,49] diminished music’s influence on cognitive outcomes, reinforcing the importance of aligning exercise intensity, music characteristics, and participant preferences to have differential effects on outcomes.

### Factors contributing to study heterogeneity

Variability in music (e.g., the use of self-selected and/or researcher-selected music) and exercise protocols emerged as a significant source of heterogeneity (see e.g., appendix tables B1 and C1). Self-selected music often aligns more closely with individual preferences and physiological rhythms, eliciting stronger emotional and motivational responses, as demonstrated by [28], where participant-selected slow-tempo music elevated affective responses by reducing mental demand during tasks. In contrast, mismatched protocols, such as slow-tempo music during high-intensity exercise, diminished both cognitive and affective outcomes [16,45]. Differences among participants not accounted for in the meta-regression analysis, such as fitness level and cultural background, may have influenced the cognitive control benefits of the music-exercise combination. For instance, individuals at higher fitness levels may be better able to sustain these cognitive benefits at higher exercise intensities by mitigating the effect of reaching VT (e.g., the point at which breathing becomes significantly more laboured during exercise) [7]. These differences influence how individuals experience music and exercise simultaneously, as affective responses, auditory-motor coupling, and tolerance for exercise intensity are integral to which cognitive resources can be directed toward such tasks.

Widely different measurement tools used across the studies contributed to heterogeneity. Tasks such as the Stroop test, while broadly used, overlap in assessing inhibitory control and attention allocation [43], introducing potential confounds and masking domain-specific effects. Latent constructs within these tools, such as mood and arousal influences, may have further magnified variability [62]. To this point, the reliability and validity of the measures used in these studies are generally well-supported, but their appropriateness for specific exercise contexts warrants scrutiny. Contextual factors such as participant fatigue or environmental conditions can compromise data reliability if the scales were not explicitly designed for such scenarios [29]. The critical issue is whether researchers selected and adequately justified instruments that were optimal for the contexts under investigation. Constructs in this body of evidence are informed by diverse theories and models—varying in factors, dimensions, and interrelationships—necessitating rigorous consideration of measurement alignment with theoretical underpinnings.

For instance, Tammen’s Attention Scale [37], developed to measure attention strategies (association and dissociation) in elite runners, may lack ecological validity when applied to other exercise contexts, potentially distorting its intended capture. Many studies relied on previously validated scales without sufficiently addressing their relevance to specific populations or contexts [48, 49]. This overreliance risks misalignment between scale design and study context. Additionally, administration methods, such as self-reports during exercise, may introduce common method biases (e.g., variance transfer) inflating variance and heterogeneity in the meta-analytic findings.

Finally, our categorisation into three meta-analytic clusters (attention allocation, inhibitory control, and core affect) may have oversimplified the analysis of the constructs measured across studies, given the specific design choices underlying these constructs. This misalignment may have contributed to the high heterogeneity observed in the meta-analyses.

## Limitations and implications

The applicability of the present review is limited to active/healthy populations. Accordingly, further examination of music and acute exercise with at-risk populations (e.g., cardiovascular disease) [63] and those that are insufficiently active, appears warranted. The high heterogeneity across the included studies and meta-analyses precludes the ability to draw definitive causal conclusions. Variability in study methodologies, including differences in exercise intensity, music protocols, outcome measurements, and participant characteristics, likely contributed to the heterogeneous results. Although extensive efforts were made to minimise publication bias through comprehensive database searches, the exclusion of conference abstracts and non-English language studies introduces the potential for language and selection biases.

Only 10 studies were included in the review, with nine eligible for meta-analysis, limiting its scope and reducing the breadth of the quantitative synthesis given the relatively small number of studies. The lack of robust longitudinal data further restricts the ability to evaluate the long-term effects of music-exercise combinations on cognitive and affective outcomes. In addition, despite large effect sizes, the wide confidence intervals and prediction intervals highlight substantial variability and uncertainty in the findings, reducing their statistical robustness.

Future research should prioritise high-quality RCTs with mediation analyses to explore the direct influence of specific musical stimuli and acute exercise protocols on cognitive control processes and affective outcomes. Such studies incorporating mediation analyses would be valuable for understanding the long-term effects, and the mechanisms through which music and acute exercise interact to influence cognitive and affective outcomes. Researchers should also strive for clearer conceptual definitions and standardised measures of cognitive control processes to facilitate the integration of findings across different studies and disciplines.

The present data indicate some utility in the adoption of low-to-moderate arousal music to match a low or moderate-intensity acute exercise protocol, and may provide a novel avenue to examine the interaction between music’s influence on recuperation from exercise. Passive recovery is intrinsic to many high-intensity exercise protocols, and further research is warranted to examine potential differences in optimal music-exercise approaches for enhancing recovery outcomes [31,64].

## Conclusion

At low-to-moderate exercise intensities, music listening during acute exercise had a differential effect on cognitive control processes and affective responses. As identified within the review, attention allocation and inhibitory control were the cognitive control processes most consistently influenced by music during acute exercise. Notably, music supported dissociative attention by diverting focus from interoceptive cues, facilitating rhythmic synchronisation, and improving inhibitory control through auditory-motor coupling. These effects were less pronounced at higher exercise intensities, where increased physiological and cognitive demands limited the influence of music. Regarding affective outcomes, music had a differential effect on core affect, particularly during low to moderate-intensity exercise, where tempo-matched music reduced fatigue perception. However, the findings are to be interpreted cautiously, due to high heterogeneity, variability in music protocol design, such as tempo mismatches or the use of researcher-selected and/or self-selected music, and individual preferences resulting in inconsistent findings across studies. Future research should prioritise high-quality RCTs with mediation analyses to explore the direct influence of specific musical stimuli and acute exercise protocols on cognitive control processes and affective responses.

Interventions incorporating low-to-moderate arousal music paired with low and moderate-intensity acute exercise protocols may provide a novel opportunity to examine the interaction between music’s influence and post-exercise recuperation.

## Data Availability

https://osf.io/xfn3m/?view_only=ba5de4d156a248998c615f24c17ef001

## Author Contributions

**Conceptualisation:** Andrew Danso, Friederike Koehler, Patti Nijhuis, Geoff Luck

**Methodology:** Andrew Danso, Keegan Knittle

**Investigation:** Andrew Danso, Julia Vigl, Joshua S. Bamford, Margarida Baltazar

**Data curation:** Andrew Danso, Julia Vigl, Joshua S. Bamford

**Formal analysis:** Andrew Danso, Keegan Knittle, Ming Yu Claudia Wong

**Writing - original Draft:** Andrew Danso, Friederike Koehler, Patti Nijhuis, Shannon E. Wright, Eero A. Haapala, Margarida Baltazar

**Writing - review & editing:** Andrew Danso, Julia Vigl, Friederike Koehler, Patti Nijhuis, Eero A. Haapala, Ming Yu Claudia Wong, Keegan Knittle, Shannon E. Wright, Nora Serres, Niels Chr. Hansen, Andrea Schiavio, Suvi Saarikallio, Geoff Luck

## Appendix

## Appendix A. Summary of operationalisation of main outcomes of interest in the review

The main outcomes of interest in the review are operationalised as follows:

- **Attention allocation** in physical exercise involves directing focus towards internal sensations (association) and/or external stimuli (dissociation) [37]. Evidence suggests that individuals allocate attention to both internal and external cues simultaneously, with the degree of focus on each type of stimuli being context-dependent [38,50].
- **Cognitive flexibility** in physical exercise refers to the dynamic interplay of attentional control and strategy adjustment, enabling individuals to effectively adapt to the evolving physical and mental demands of exercise. This involves the ability to shift focus between internal and external stimuli, modify exercise strategies based on feedback and changing conditions, and regulate cognitive processes [1].
- **Core affect** is the fundamental experience of feeling, encompassing both emotions and moods [29]. It is characterised by two core dimensions: valence (pleasure-displeasure) and activation (arousal-sleepiness). Affect arises from a complex interplay of physiological processes, cognitive appraisals, and situational influences, and its expression can range from basic, reflexive responses to complex, nuanced emotions.
- **Inhibitory control**, crucial for suppressing impulsive actions (such as abrupt changes in intensity or technique) and resisting distractions from both internal (e.g., negative thoughts) and external sources (e.g., environmental factors), is essential for regulating attention and making deliberate decisions during physical exercise [1].
- **Task switching**, a component of cognitive flexibility, involves the coordinated interplay of attentional functions, including interference suppression, to enable shifts between different cognitive and motor skills [1]. The efficiency and speed of task switching are influenced by factors like task complexity, individual differences, and practice, and play a crucial role in adapting to the dynamic demands of physical exercise.
- **Working memory**, a system that actively holds, manipulates, and processes information, is essential for retaining and applying critical information during physical exercise, including instructions, goals, pacing, and technique [39]. It enables individuals to monitor physiological feedback, perform mental calculations, plan and execute movement sequences, and adapt to dynamic exercise demands. Working memory shares neural mechanisms with attention, relying on interconnected brain regions such as the prefrontal and parietal cortex to support cognitive control during physical exercise [39].

## Appendix B. Summary of findings of Randomised Controlled Trials

Two RCTs [45,48] investigated the effect of music during acute exercise on various cognitive processes and affective outcomes, examining outcomes in inhibitory control , working memory and core affect. Across the trials, music used during exercise was consistently associated with improvements in inhibitory control and working memory (measured post-exercise) compared to exercise without music. [45] measured inhibitory control immediately after a 20-minute, moderate-intensity aerobic exercise session. [48] measured inhibitory control using the Stroop test within 30 seconds of exercise cessation, with all participants finishing within three minutes (see e.g., Table B1 for a summary of the studies).

**Table B1.**
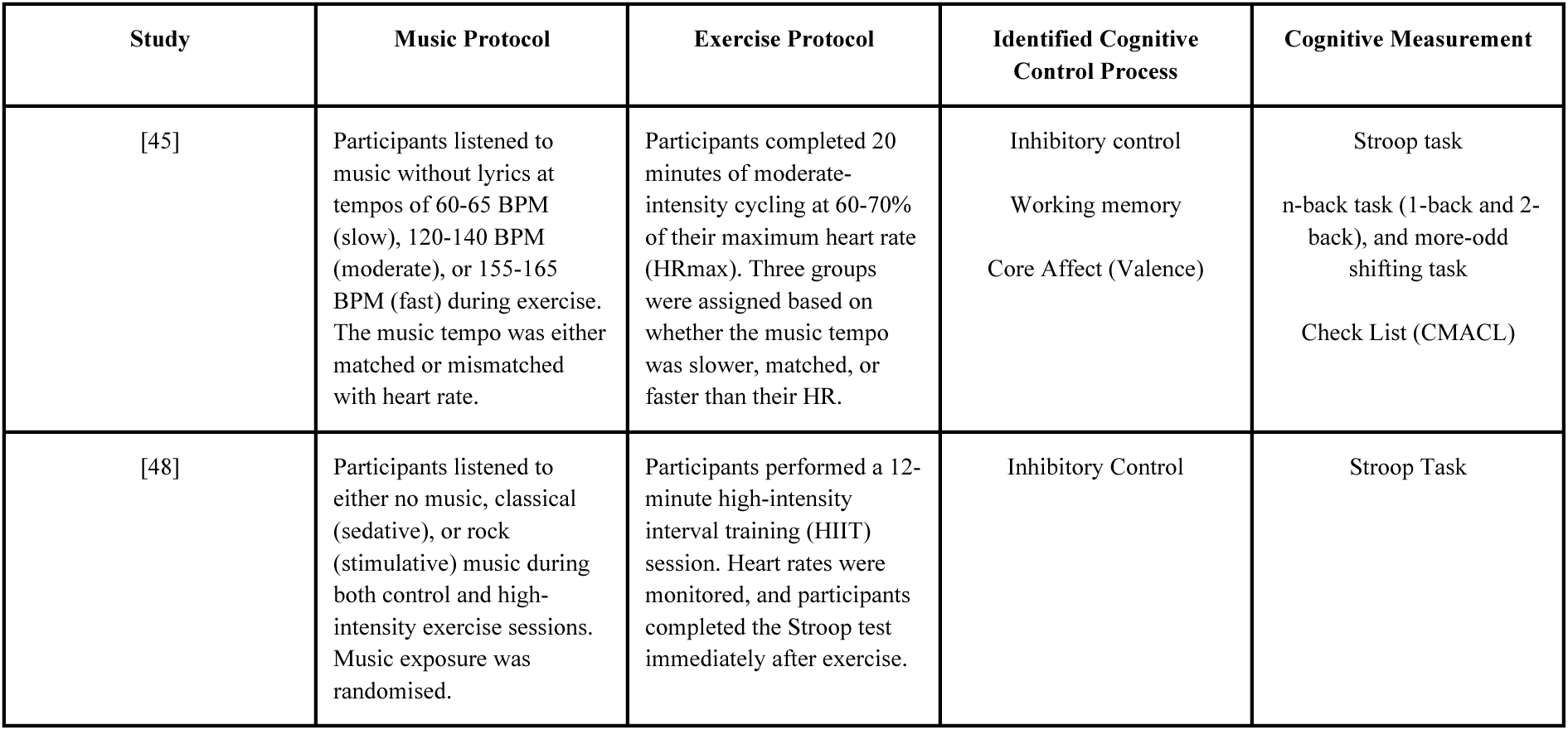
Summary table of included studies that were RCTs.

## Appendix C. Summary of findings of experimental studies

Eight studies used a variety of experimental designs to examine the effects of music listening during acute exercise on cognitive control processes and affective outcomes, reporting outcomes across attention allocation [13,14,46,49], core affect [13,14,46,49,51], inhibitory control [44,51], and working memory [52] (see e.g., Table C1 for a summary of the studies).

**Table C1.** Summary table of included studies with other experimental designs than RCTs.

| Study | Music Protocol | Exercise Protocol | Identified Cognitive Control Process | Cognitive Measurement |
| --- | --- | --- | --- | --- |
| [14] | Participants listened to the song “I Heard It Through the Grapevine” (119 BPM) by Creedence Clearwater Revival during a 10-minute isometric handgrip exercise session. The music was delivered through MRI-compatible earphones at a sound intensity of ~75 dBA. | Participants performed 30 trials of a 10-second isometric handgrip exercise at 30% of their maximal voluntary contraction (MVC), followed by 10 seconds of rest. The exercise was performed while inside an MRI scanner to measure brain activity during the task. | Attention Allocation<br>Core Affect (Arousal) | Attentional Scale (Tammen’s scale)<br>Felt Arousal Scale |
| [13] | Participants listened to the song “Happy” (160 BPM) by Pharrell Williams during a 6-minute outdoor walking session on a 400-meter running track at a self-paced speed. | Participants walking 400 meters at a self-paced speed on a standard outdoor running track. Each participant completed this walking task under three different conditions: music, podcast, and control (no auditory stimulus). The auditory stimuli were randomised and counterbalanced across participants. | Attention Allocation<br>Core Affect (Arousal) | Attentional Scale (Tammen’s scale)<br>Felt Arousal Scale |
| [44] | Participants listened to researcher-chosen, high-decibel music at 100 dB, considered high-intensity sound (HIS), during exercise sessions. | Participants cycled on an ergometer at 70-75% of VO<sub>2</sub> max (a level that constitutes high-intensity exercise). | Inhibitory Control | Stroop Task |
| [46] | Participants listened to either fast-tempo music (120 BPM) or slow-tempo music (90 BPM), with tracks selected from the Billboard Hot 100 Chart. The music was played throughout the duration of the exercise at a constant volume of 75 dBA using external speakers. The fast | Participants performed two isometric strength exercises: a wall-sit and a plank-hold. In the wall-sit, participants maintained a seated position with their back against the wall and knees at a 90-degree angle, holding the position until voluntary exhaustion. For the plank-hold, participants held a | Attention Allocation<br>Core Affect (Arousal & Valence) | Attentional Scale (Tammen’s scale)<br>Affect Grid |
|  | and slow-tempo tracks were edited to maintain consistent tempos, ensuring the differentiation between music conditions. A no-music control condition was also used for comparison. | prone plank position, supported by their elbows and feet, until they could no longer maintain proper form. Both exercises were completed in a baseline (no-music) trial, followed by an experimental trial with either no music, fast-tempo music, or slow-tempo music. |  |  |
| [49] | Participants completed three conditions: self-selected music (high-tempo favorite tracks), randomly selected music (a "Sport" playlist), and no music. Music (140-160 BPM) played during the 5-minute warm-up, exercise bouts, and recovery periods via headphones, with volume standardised at 75%. Music's motivational properties were evaluated post-session. | The sprint interval training (SIT) involved 8 × 15-second all-out cycling bouts at 9% body mass resistance, with 120 seconds of passive recovery between bouts. Performance metrics (peak power, mean power, fatigue index, and total work) were recorded, and no verbal encouragement was provided. Sessions were spaced 72 hours apart for recovery. | Attention Allocation<br>Core Affect (Valence) | Attentional Scale (Tammen's scale)<br>Feeling Scale |
| [51] | Participants listened to their favourite music or metronome beeps during 10 minutes of moderate-intensity pedalling exercise. The music had a tempo of 120 beats per minute, synchronised with the pedalling. | Participants performed 10 minutes of moderate-intensity pedalling exercise (50% of their peak oxygen uptake) on a recumbent cycle ergometer at 60 revolutions per minute. | Inhibitory Control<br>Core Affect (Valence) | CWST<br>Two-Dimensional Mood Scale (TDMS) |
| [16] | Participants listened to self-selected music through headphones during the exercise. Songs were Japanese pop music with a tempo below 120 BPM. The volume was set to 80 dB. | Participants performed 30 minutes of moderate-intensity cycling at 60% VO<sub>2</sub> peak. They maintained a cadence of 60 revolutions per minute. | Inhibitory Control<br>Core Affect (Arousal) | Stroop Task<br>Felt Arousal Scale |
| [52] | Participants listened to a 30-minute classical music playlist (120-140 BPM) with noise-cancelling headphones at a consistent volume during the session. | Participants exercised on a recumbent cycle ergometer for 30 minutes at a moderate intensity (55% of their heart rate reserve), including a 5-minute warm-up and cool-down. | Inhibitory Control<br>Working Memory | Stroop Task<br>Reverse Corsi Block Task |

## Appendix D. Publication bias: sensitivity analysis

A sensitivity analysis was conducted to assess publication bias by relating standard errors to effect size estimates across the outcomes of interest. Following the recommendations of [65,66], a series of funnel plots of per-outcome standard error by standard difference in group means was produced and assessed for evidence of asymmetry. Forest plots summarised the effect size data, while funnel plots were used to explore potential publication bias.

Egger’s test [65] indicated significant asymmetry for attention allocation (*z* = 2.953, *p* = .003), significant asymmetry for inhibitory control (*z* = 4.796, *p* < .001), and significant asymmetry for core affect outcomes (*z* = 3.275, *p* = .001) (Figure D1). Because of potential publication bias, the summary effect sizes for attention allocation, inhibitory control and core affect outcomes may thus be slightly inflated.

**Figure D1.**
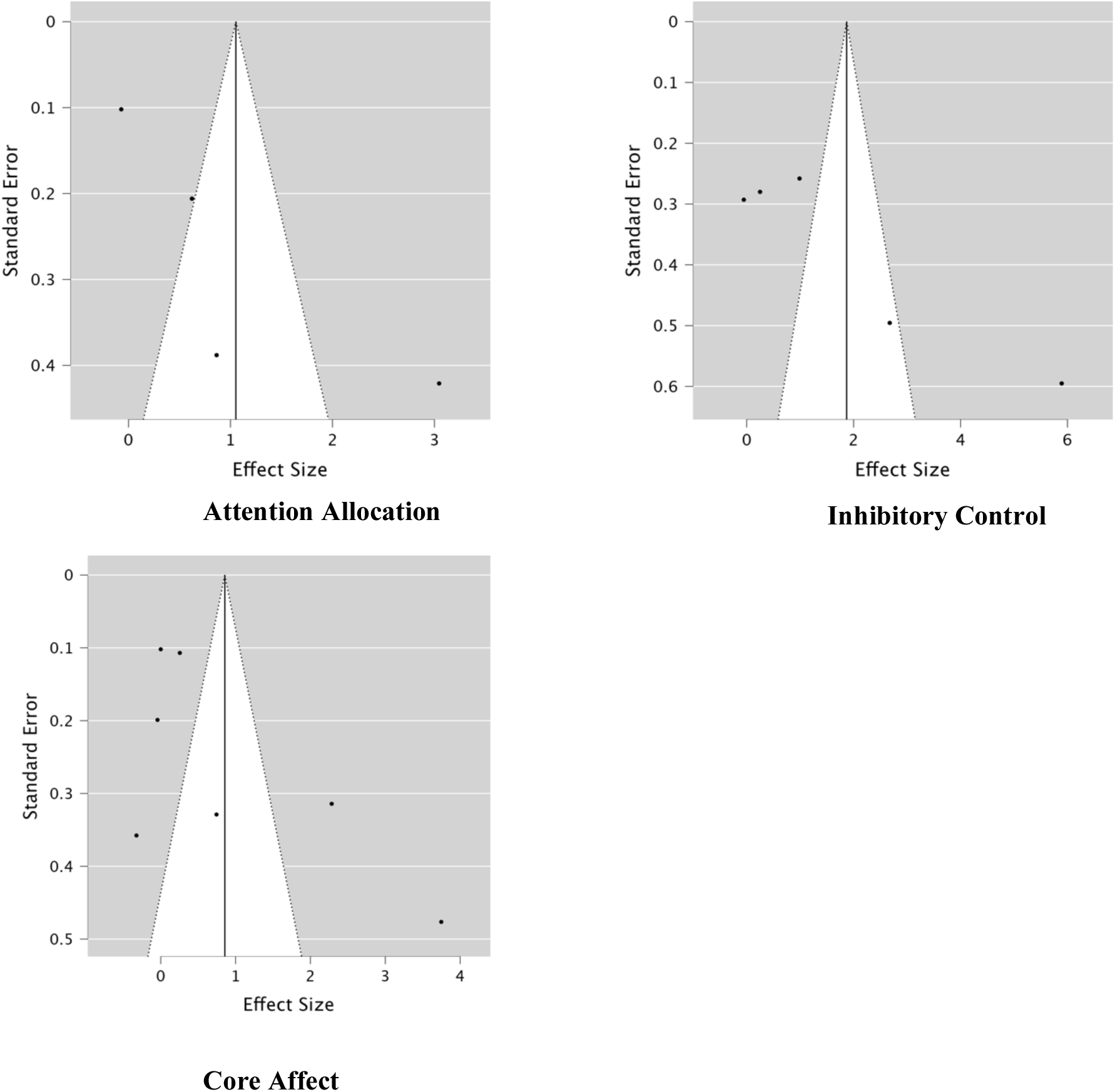
Funnel plots for attention allocation, inhibitory control and core affect outcomes.

